# Integration of Gene Expression and Digital Histology to Predict Treatment-Specific Responses in Breast Cancer

**DOI:** 10.1101/2025.08.25.25334393

**Authors:** Frederick M. Howard, James Dolezal, Hanna Hieromnimon, Sara Venters, Sara Kochanny, Anran Li, Alexander Borowsky, W. Fraser Symmans, Denise Wolf, Lamorna Brown-Swigart, Anthony Sun, Amrita Basu, Gillian Hirst, Long C Nguyen, Adam Asare, Sai Kanaparthi, Galina Khramtsova, Kim Blenman, Naing Lin Shan, Cheng Fan, Sara M. Tolaney, George Somlo, Clifford A. Hudis, William Sikov, Linda McCart, Mark Watson, Lisa Carey, Daniel G. Stover, Laura Van’t Veer, Laura J Esserman, Charles M. Perou, Lajos Pusztai, Olofunmilayo I. Olopade, Dezheng Huo, Rita Nanda, Alexander T. Pearson

**Affiliations:** Department of Medicine, University of Chicago, Chicago, IL, USA; Geisinger Cancer Institute, Danville, PA, USA; UCSF Helen Diller Family Comprehensive Cancer Center, University of California, San Francisco, CA, USA; Department of Pathology and Laboratory Medicine, University of California, Davis, CA, USA; Department of Pathology, The University of Texas MD Anderson Cancer Center, Houston, TX, USA; Quantum Leap Healthcare Collaborative, San Francisco, CA, USA; Department of Medicine, Yale School of Medicine, New Haven, CT, USA; Lineberger Comprehensive Cancer Center, University of North Carolina at Chapel Hill, Chapel Hill, NC, USA; Dana-Farber/Harvard Cancer Center, Boston, MA, USA; City of Hope Comprehensive Cancer Center, Duarte, CA, USA; Memorial Sloan Kettering Cancer Center, New York, New York; Program in Women’s Oncology, Women and Infants Hospital of Rhode Island, Warren Alpert Medical School of Brown University, Providence, RI, USA; The Ohio State University Comprehensive Cancer Center, Columbus, OH; Department of Pathology and Immunology, Washington University School of Medicine, St. Louis, MO, USA; Department of Public Health Sciences, University of Chicago, Chicago, IL, USA

## Abstract

Deep learning models applied to digital histology can predict gene expression signatures (GES) and offer a low-cost, rapidly available alternative to molecular testing at the time of diagnosis. We optimized transformer-based models to infer GES results and applied this approach to pre-treatment H&E-stained biopsies from 1,940 breast cancer patients treated with neoadjuvant chemotherapy in clinical trial and real-world cohorts. The most predictive histology-derived GES for pathologic complete response (pCR) in the I-SPY2 trial was validated in four external cohorts: CALGB 40601, CALGB 40603, a trial of durvalumab plus CT, and standard-of-care CT-treated patients from the University of Chicago. Among HER2-negative patients, a transformer-based model trained using a signature composed of estrogen-regulated genes, proliferation, apoptosis, and interferon response genes predicted pCR with an AUC of 0.794, outperforming models based on clinical features alone (AUC 0.704, p = 0.001), pathologist TIL assessment, and a model trained directly to predict response from I-SPY2 cases. Tertiles of this signature stratify patients into clinically relevant groups with increasing likelihood of complete response, with pCR rates ≥50% in the top tertile regardless of treatment or hormone receptor status. Additional transformer-based signature models predicted response to specific therapies (but not chemotherapy alone), including a HER2 signaling signature in IO-treated patients, and a claudin-low signature in bevacizumab treated patients. In HER2- cohorts with available gene expression data and histology, models trained on expression data performed similarly to digital histology predictions, but the combination of gene expression and histology outperformed histology alone. These findings suggest that histology-based GES provides additive information to RNA sequencing data and can inform precision treatment selection across breast cancer subtypes.

## Background

Breast cancer remains one of the most heterogeneous malignancies, posing a challenge to both prognostication and therapeutic decision-making. Gene expression profiling has played a key role in understanding breast cancer heterogeneity, revealing distinct molecular subtypes to inform risk of recurrence and response to therapy^1–3^. However, worldwide adoption of transcriptomic testing is limited by factors including tissue availability, assay cost, and formalin fixation degrading RNA integrity. In parallel, there has been a rapid expansion of computational pathology approaches to predict clinically actionable biomarkers from hematoxylin and eosin (H&E)-stained slides^4,5^ – which are routinely available for all patients with cancer diagnosis. Historically, research groups have demonstrated that certain morphological features correlate with the expression of specific genes associated with proliferation, immune infiltration, hypoxia, and other pathways crucial to cancer progression. Indeed several immune-related gene signatures – for instance, those capturing activated CD4+ T-cell activity or cytokine signaling – show high biological and clinical relevance in breast cancer^6^. Similarly, estrogen-regulated signatures have been integral to understanding hormone receptor (HR)–positive disease biology^7^. Improvements in computational methodologies offer the opportunity to directly infer these and hundreds of other gene signatures from histologic images, potentially streamlining biomarker discovery and facilitating precision oncology strategies.

Multiple prior studies have successfully reconstructed bulk gene expression from pathology slides^8,9^, and leveraged predicted gene expression to predict response to therapy and long term outcomes in cancer. Despite these advances, accurately predicting the phenotypic impact of thousands of individual gene transcripts remains a formidable challenge. The past year has seen rapid growth in histology-specific foundational models^10,11^ – which can better capture distinct features from digital pathology images; whereas these older approaches to predicting transcriptomics rely heavily on general image analysis models. One key difficulty is handling the vast spatial resolution of whole-slide images, which can contain millions of pixels. Multiple instance learning (MIL) has emerged as a powerful framework to address this problem^12^. By dividing a slide into smaller tiles (instances) and assigning only a slide-level label, MIL allows a model to learn which tiles are most salient for a given outcome or feature of interest. More recently, transformer-based approaches have adapted MIL by incorporating attention mechanisms^13,14^, enabling the model to selectively weight informative tiles and potentially capture more intricate spatial relationships that might correlate with gene expression programs. Finally, although these approaches have demonstrated some accuracy in predicting gene expression, the correlation between true / predicted gene expression remains poor for the average gene, and thus it is challenging to draw conclusions about the biologic basis of predictions. Conversely, gene signatures generally exhibit higher biological robustness and reproducibility; their interpretability can be more straightforward, as they represent well-defined pathways or cell populations; and gene signatures can be predicted with much greater accuracy than the expression levels of individual genes^15^.

We therefore sought to create and optimize a Transformer-based hIstology-driven Gene Expression Regressor (TIGER) from H&E slides, focusing on a comprehensive set of 775 biologically validated breast cancer signatures. These signatures were previously curated to encapsulate core processes in breast tumorigenesis, immune response, hormone signaling, and more. We applied this model to predict response to neoadjuvant therapy in breast cancer, as there are numerous gene expression assays that are either commercially available or in development to predict response to therapy for all subtypes of breast cancer^16–19^, and a digital histology model for expression signatures has the potential to provide similar information without the cost and tissue consumption of traditional assays. We also evaluate the ability of histology-derived gene expression to identify specific biomarkers of immunotherapy response and response to other targeted therapies, as standard biomarkers such as PD-L1 status fail to identify patients who benefit from immunotherapy with early-stage HER2- breast cancer^20^.

## Results

### Optimizing a Pipeline for Gene Expression Signature Prediction

We first sought to develop an optimized pipeline for gene expression and gene signature prediction, using data from TCGA and CPTAC, across six cancer subsets (BRCA, COAD, LUAD, LUSC, GBM, and HNSC). We optimized hyperparameters including foundation model, feature aggregation and learning rate using five-fold cross-validation in TCGA to predict bulk gene expression as well as 775 breast cancer gene expression signatures (Supplemental Table 1), resulting in a pipeline we term Transformer-based hIstology-driven Gene Expression Regressor (TIGER). This pipeline was also compared to state-of-the-art methods of gene expression prediction (**Figure 1B**, **Supplemental Table 1**) – including SEQUOIA and DeepPT (both using the same input foundational model features to allow for a consistent comparison). TIGER outperformed these alternative methods for bulk gene expression prediction in internal five-fold cross validation in TCGA and external testing in CPTAC for all cancer types / cohorts except the HNSC CPTAC cohort (**Supplemental Table 2**).

**Figure 1.**
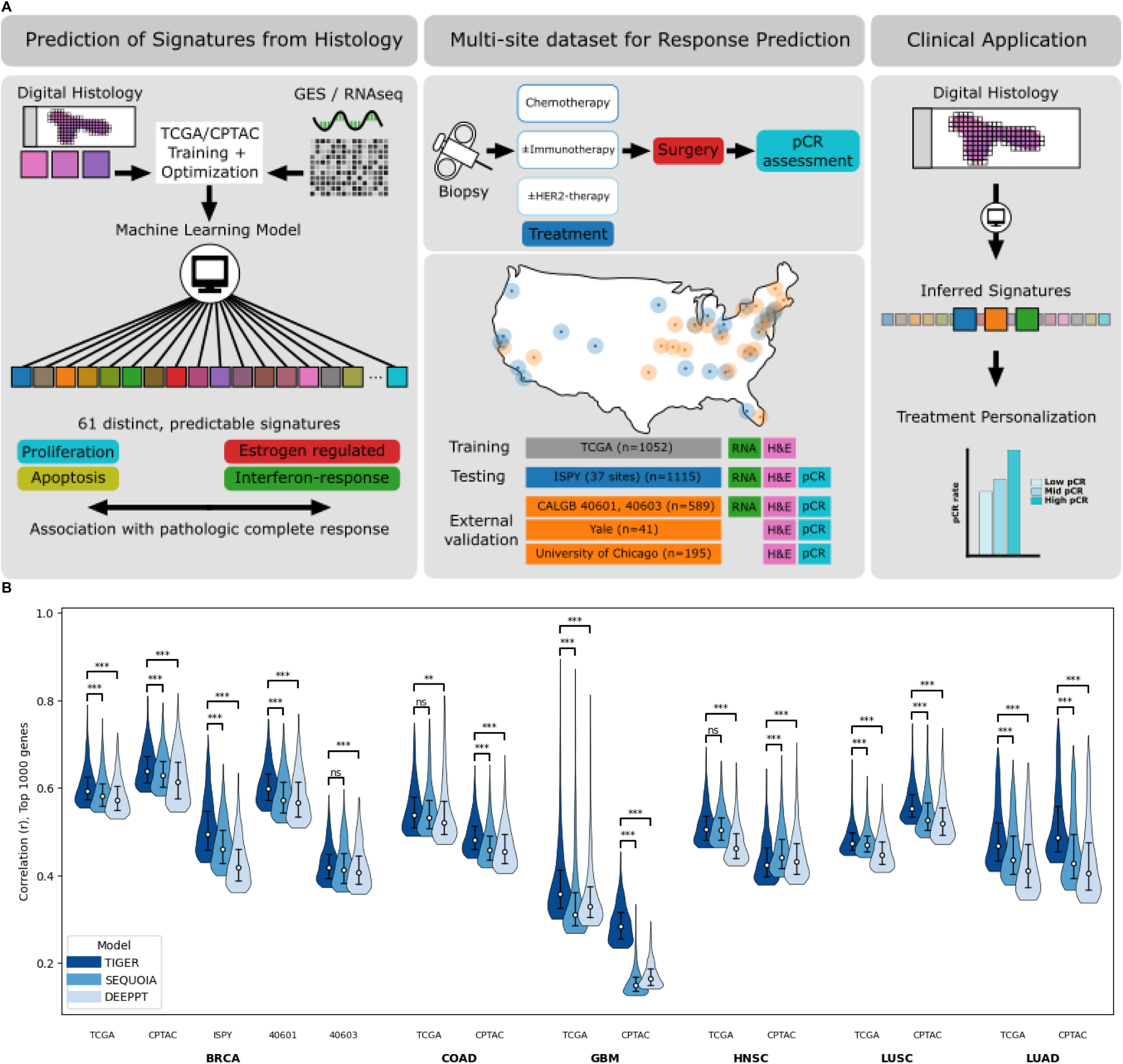
Clinically Actionable Gene Expression and Gene Signature Prediction. **A.** Study Overview. We designed a transformer-based model that accurately predicts bulk gene expression as well as a wide array of distinct gene expression signatures from digital histology. We then curated a dataset of over 4,000 slides from 1,400 patients receiving neoadjuvant therapy from four clinical trials and one real world cohort. Association of all predictable signatures top signatures for 1) response prediction in HER2- patients, 2) response prediction in HER2+ patients, and 3) specific response prediction to immunotherapy were identified using the I-SPY2 trial cohort. These signatures were then additionally validated in the other independent datasets. **B**. We compared the predictive accuracy of our approach for gene expression prediction pipeline, transformer-based hIstology-driven Gene Expression Regressor (TIGER), to other state of the arts methods (SEQUOIA and DeepPT). Models were trained and internally validated with five-fold cross validation in TCGA; and externally tested in CPTAC, and for the BRCA cohort, also externally tested in I-SPY2, CALGB 40601, and CALGB 40603. Visualized is the distribution of Pearson correlation coefficients of the top 1,000 predicted genes in each cohort. *** indicates p < 0.001, ** indicates p < 0.05, ns indicates not significant. **Abbreviations: BRCA**, breast invasive carcinoma; **COAD**, colon adenocarcinoma; **GBM**, glioblastoma multiforme; **HNSC**, head and neck squamous cell carcinoma; **LUAD**, lung adenocarcinoma; **LUSC**, lung squamous cell carcinoma; **TCGA**, The Cancer Genome Atlas; **CPTAC**, Clinical Proteomic Tumor Analysis Consortium; **I-SPY2**, Investigation of Serial Studies to Predict Your Therapeutic Response with Imaging and Molecular Analysis 2; **CALGB**, Cancer and Leukemia Group B; **HER2**, human epidermal growth factor receptor 2; **TIGER**, hIstology-driven Gene Expression Regressor; **SEQUOIA**, Spatially Enabled Quantification Of Individualized Alterations; **DeepPT**, Deep learning-based Prediction of Transcriptomics.

The BRCA models were also tested in the I-SPY, CALGB 40601, and CALGB 40603 clinical trial cohorts on pre-treatment biopsies where gene expression data was available. Of note, TIGER achieved the best performance in all breast cancer datasets, as measured by correlation coefficient of true vs predicted gene expression of the top 1,000 genes predicted per model per dataset (mean *r* 0.604 in TCGA, 0.647 in CPTAC, 0.509 in I-SPY, 0.607 in CALGB 40601, and 0.427 in CALGB 40603), outperforming SEQUOIA (mean *r* 0.591 in TCGA, 0.637 in CPTAC, 0.470 in I-SPY, 0.583 in CALGB 40601, and 0.423 in CALGB 40603) and DeepPT (mean *r* 0.581 in TCGA, 0.623 in CPTAC, 0.430 in I-SPY, 0.579 in CALGB 40601, and 0.419 in CALGB 40603) – all comparisons significant except TIGER vs SEQUOIA in CALGB 40603 (p = 0.060).

We applied TIGER to predict the continuous values of 775 pre-determined gene signatures from TCGA-BRCA (n = 1,052 cases). The average signature was predicted with a median correlation coefficient of 0.54 (range 0.11 – 0.85, **Figure 2A**). Predictions across the 775 signatures generally clustered into groups related to proliferation / basal biology, luminal biology, immune infiltrate, and vascular/hypoxia signaling. Signatures from the luminal group were most accurately predicted from histology, with the highest accuracy seen for a Luminal A signature with a correlation coefficient for predicted vs. true signature of 0.83 (Garcia-Recio, JCI 2020)^21^. Signatures were accurately predicted in external cohorts including CALGB 40601 (n = 129, median correlation coefficient 0.41) and CALGB 40603 (n = 195, median correlation coefficient 0.25) – of note, in these cohorts, predictions were made on pretreatment biopsies and for single cancer subtypes with lower variance in subtype-specific signatures (**Figure 2B**). The hierarchy and predictability of signature clusters were further visualized with a UMAP representation (**Figure 2C**). Interestingly, the histology-derived IIE signature (related to estrogen related genes) clustered with immune gene signatures but also appeared to be closely associated with basal / proliferation signatures. Top 25 predicted signatures fell into two anti-correlated groups of luminal signatures and proliferation/basal signatures (**Figure 2D**), all with correlation of > 0.75 versus the true signature in TCGA; with a median correlation coefficient of 0.64 in CALGB 40601 and median correlation coefficient of 0.40 in CALGB 40603 among these top signatures. In the I-SPY2 cohort, where a mix of FFPE and frozen biopsy cores were taken from a subset of patients, the consistency of signature predictions was also evaluated. The intraclass correlation coefficient was high for the set of 775 signatures when computed across a pair of FFPE slides (**Figure 2E**), with a median of 0.84; many signatures were also similar when computed from FFPE versus frozen slides from the same patient (**Figure 2F**), with a median intraclass correlation coefficient of 0.72. A scatter plot of the correlation between predictions from two slides from the same patient for a select signature (the above-mentioned IIE signature) further illustrates the high consistency for these predictions (**Figure 2G**).

**Figure 2.**
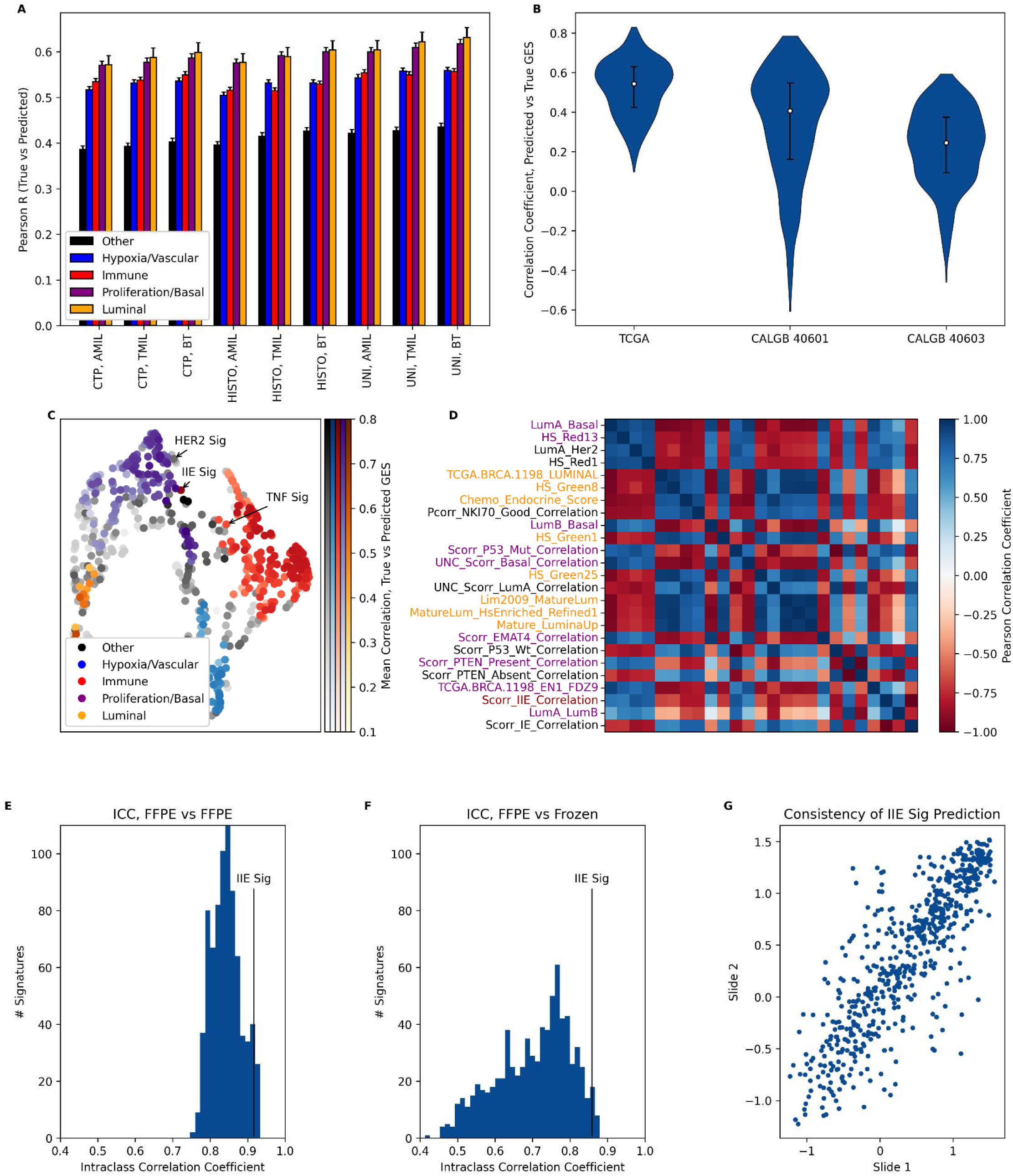
Exploring Gene Expression Signature Prediction from Digital Histology. **A.** We optimized approaches to predict gene expression signatures from digital histology. Performance for gene signature prediction on fivefold cross validation in TCGA is shown. We evaluated different foundational models, including CTransPath (CTP), HistoSSL (HISTO), and UNI, as well as approaches to feature aggregation including multiple-instance learning (MIL) models such as attention MIL (AMIL) and TransMIL (TMIL), as well as the Bistro Transformer (BT). The optimal combination (UNI + BT) forms the basis for our transformer-based hIstology-driven Gene Expression Regressor (TIGER) approach for signature prediction. **B.** Accuracy of our optimized approach in the TCGA training cohort, as well as the CALGB 40601 and CALGB 40603 validation cohorts for gene expression signature prediction. **C.** The predicted gene expression signatures from digital histology form unique clusters of correlated signatures, predominantly related to immune genes, proliferation, luminal biology, and hypoxia signals. The intensity of shading indicates the accuracy of prediction of each signature on fivefold cross validation in TCGA. **D**. Shown is a correlation matrix of the 25 most accurately predictable signatures. As shown, these signatures form two anti-correlated groups, representing luminal / low grade and basal / high grade tumors. **E.** A histogram of the **i**ntraclass correlation coefficient for model predictions of all 775 signatures, taken from two FFPE slides among patients (n = 601) with multiple FFPE slides in the I-SPY2 cohort is shown. **F.** Intraclass correlation is shown for signature prediction from FFPE versus fresh frozen tissue specimens among patients (n = 141) with both specimen types in the I-SPY2 cohort. **G.** A scatter plot illustrating the consistency of predictions for the IIE signature in the I-SPY2 dataset across two separate FFPE slides, among patients with multiple slides. **Abbreviations**: **AMIL**, attention-based multiple-instance learning; **BT**, Bistro Transformer; **CALGB**, Cancer and Leukemia Group B; **CTP**, CTransPath; **FFPE**, formalin-fixed, paraffin-embedded; **HISTO**, HistoSSL; **I-SPY2**, Investigation of Serial Studies to Predict Your Therapeutic Response with Imaging and Molecular Analysis 2; **MIL**, multiple-instance learning; **TCGA**, The Cancer Genome Atlas; **TIGER**, hIstology-driven Gene Expression Regressor; **TMIL**, TransMIL.

### Application for Response Prediction in HER2- Breast Cancer

After exclusion of patients without digital images, absent response data, and nonstandard treatments (**Extended Data** Figure 1), we obtained 5,293 slide images from 1,940 patients across four clinical trials and one real world patient cohort (**Supplemental Table 3, Supplemental Table 4**) for association of predicted signatures with response. This included 1,115 patients from the I-SPY2 clinical trial, 359 and 230 patients from Alliance trials CALGB 40603 and 40601, 41 patients from a single arm trial of durvalumab + neoadjuvant chemotherapy, and 195 patients receiving standard of care therapy at University of Chicago. This cohort includes predominantly HER2 negative disease (n = 1,280, 74.2%); 868 (50.3%) had HR negative disease, and 621 (36.0%) had pathologic complete response to neoadjuvant therapy.

Digital images were extracted using automated quality control with Otsu thresholding and Gaussian blur-based filtering resulting in a similar proportion of usable tiles across datasets (**Extended Data** Figure 2A-C). Predictive accuracy of histology-derived signatures was first formally assessed in HER2- FFPE samples from the I-SPY2 cohort (n = 579, **Figure 3A**) – given the larger number of samples available in this cohort. Statistical testing was performed after removing poorly predictable signatures (correlation coefficient < 0.5 versus true signature, **Figure 3B**) and co-correlated signatures (**Figure 3C**), resulting in 61 distinct signatures for evaluation (**Extended Data** Figure 3, **Supplemental Table 5**). Of note, immune signatures such as an activated CD4^+^ cell signature^22^ and a TNFα signature^23^ had high predictive accuracy for response to neoadjuvant therapy. However, a signature (termed IIE) that was originally defined using an estrogen regulated gene set analysis of ER+ breast cancers^24^ – and includes many proliferation, apoptosis, and interferon-response genes – had the greatest accuracy for response prediction, with an average AUROC of 0.788 (95% CI 0.748 - 0.829, p < 0.001) across all HER2- patients in I-SPY2. Predictive accuracy was maintained across individual arms in I-SPY2, in hormone receptor positive (AUC 0.815, 95% CI 0.762 - 0.868, p < 0.001) and negative (AUC 0.705, 95% CI 0.642 - 0.767, p < 0.001) patients, and in the UChicago (AUC 0.757, 95% CI 0.673 – 0.841, p < 0.001), Yale (AUC 0.737, 95% CI 0.578 - 0.895, p = 0.005) and CALGB 40603 (AUC 0.652, 95% CI 0.577 - 0.728) datasets (**Figure 3D**). Although the IIE signature was chosen for formal assessment of response prediction, we found that there was a high degree of consistency in the predictive accuracy of most signatures in I-SPY2 and these additional external cohorts (**Extended Data** Figure 4). To further support the use of histology gene signatures in place of true gene expression data, we compared the predictive accuracy of the 61 histology-derived gene signatures and the true value of these gene expression signatures (as measured by AUROC for pathologic complete response). Predictions from histology versus gene expression were moderately correlated in the CALGB 40601 (r = 0.49) and CALGB 40603 (r = 0.43) cohorts where these signatures have previously been computed (**Extended Data** Figure 5).

**Figure 3.**
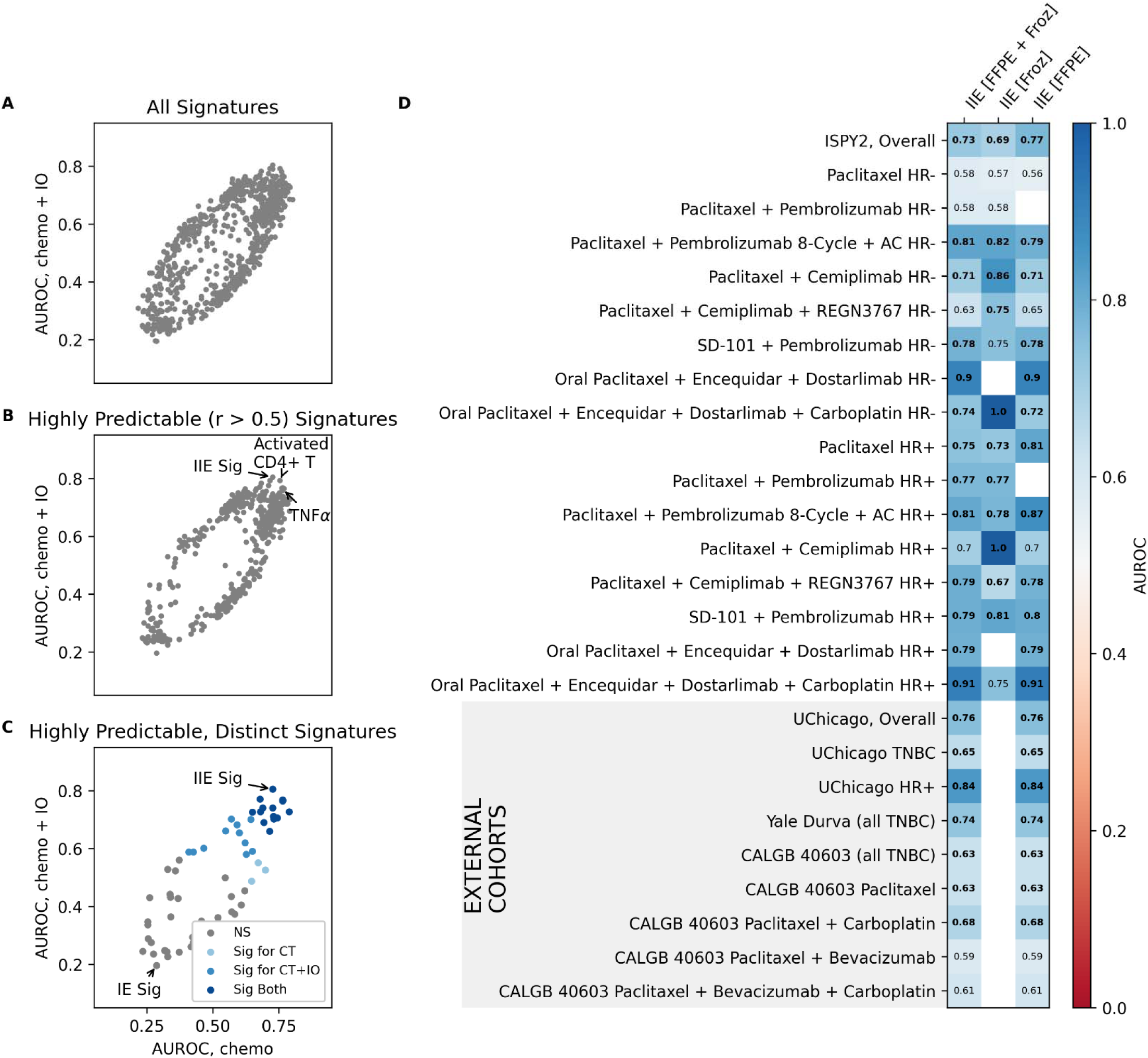
Signature Prediction for General Response Prediction in HER2- Patients. A,B,C. Shown the association of predicted signatures with pCR in FFPE samples from HER2- patients in I- SPY2. All 775 signatures are shown in **A**, with only predictable signatures (with correlation coefficient > 0.5) shown in **B**, and only distinct signatures (i.e. not co-correlated) shown in **C** where statistical testing with FDR correction was performed. The IIE signature, representing expression of genes regulated by the estrogen receptor including proliferation and immune-related genes, was most associated with response on average across HER2- arms in I-SPY2 – with other top signatures associated with response generally related to immune gene expression (as shown in B). A HER2 signature (shown in C) was associated with response to chemotherapy + immunotherapy but not chemotherapy. **D.** Shown is the association of the IIE signature with response across treatment arms, by hormone receptor status, and in frozen versus FFPE specimens, as measured by AUROC for pCR. **Abbreviations: AC**, anthracycline + cyclophosphamide; **AUROC**, area under the receiver operatingcharacteristic curve; **CALGB**, Cancer and Leukemia Group B; **CT**, chemotherapy; **ER**, estrogen receptor; **FFPE**, formalin-fixed, paraffin-embedded; **FDR**, false discovery rate; **Froz**, fresh-frozen specimen; **HER2**, human epidermal growth factor receptor 2; **HR**, hormone receptor; **IO**, immunotherapy; **I-SPY2**, Investigation of Serial Studies to Predict Your Therapeutic Response with Imaging and Molecular Analysis 2; **NS**, not significant; **pCR**, pathological complete response; **TNBC**, triple-negative breast cancer; **TNF**α, tumor necrosis factor alpha.

The IIE signature was more predictive than a logistic regression on clinical variables alone in the full I-SPY2 dataset (AUC 0.794 versus AUC 0.704, p = 0.001, **Figure 4A**), but combination of clinical variables with the gene signature did not improve predictive accuracy (AUC 0.805 versus AUC 0.794, p = 0.12). Additionally, a model trained from scratch from I-SPY2 FFPE slides using five fold cross validation to predict pCR, using the same architecture as TIGER (mean AUC 0.780), did not outperform the histology-predicted IIE signature. Pathologist TIL annotations were available from 516 patients in external cohorts (including both HER2- and HER2+ disease) – the IIE signature outperformed pathologist TIL calculations both overall (AUC 0.689 versus AUC 0.627, p = 0.005) and in each individual cohort (**Figure 4B, Supplemental Table 6**). The IIE signature prediction distribution was similar across included cohorts as well as in frozen vs FFPE samples, but was higher in TNBC samples and higher values were seen with lower numerical RCB scores (**Figure 4C**).

**Figure 4.**
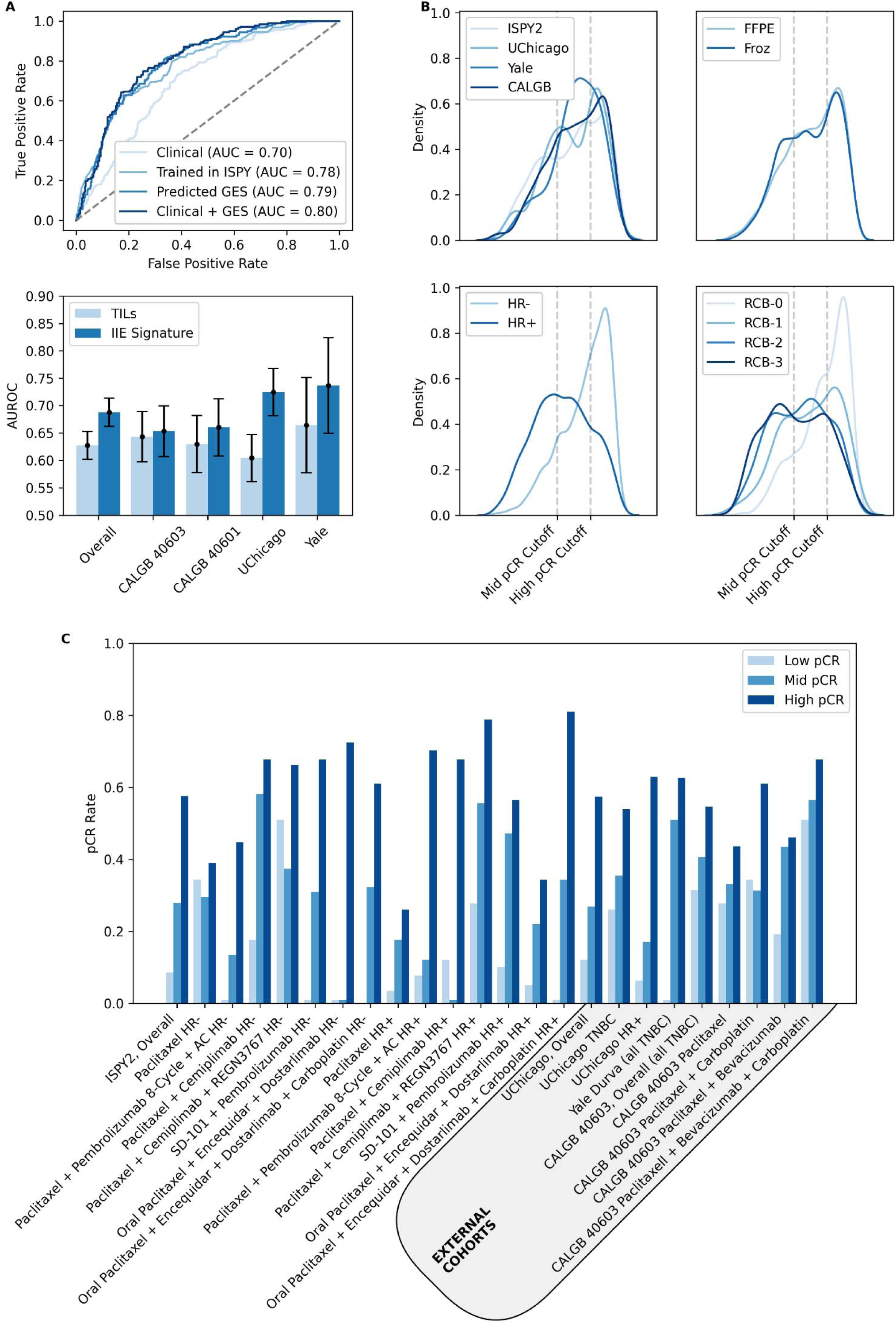
A Clinically Applicable Biomarker for Response Prediction. **A.** The predicted IIE gene signature was compared to a logistic regression on clinical variables alone in the I-SPY2 cohort, the performance of an AI model trained from scratch in I-SPY2 (with average performance on 5-fold cross validation plotted here), as well as to a combination of clinical variables and the IIE signature. **B.** Comparison of the AUROC for pathologic complete response of pathologist TIL annotations and IIE signature predictions across cohorts where annotations were available. **C.** Distribution of IIE signature predictions across relevant subgroups – including across the individual neoadjuvant cohorts of the study, the type of tissue, hormone receptor status, and residual cancer burden. **C.** Pathologic complete response rates in subgroups defined by tertiles of signature prediction (calculated from the full I-SPY2 HER2- cohort and applied to all cohorts / subgroups) – across treatment arms and by hormone receptor status. **Abbreviations: AUROC**, area under the receiver operating characteristic curve; **CALGB**, Cancer and Leukemia Group B; **FFPE**, formalin-fixed, paraffin-embedded; **Froz**, fresh-frozen specimen; **GES**, gene expression signature; **HER2**, human epidermal growth factor receptor 2; **HR**, hormone receptor; **I-SPY2**, Investigation of Serial Studies to Predict Your Therapeutic Response with Imaging and Molecular Analysis 2; **pCR**, pathological complete response; **RCB**, residual cancer burden; **TIL**, tumor-infiltrating lymphocyte; **TNBC**, triple-negative breast cancer.

Tertiles of the signature in HER2- patients defined subgroups with clinically meaningful differences in pCR rate across treatment arms - the pCR rate was consistently highest in the top tertile and was ≥ 50% in all external cohorts including HR+ and HR- cases (**Figure 4D**, **Supplemental Table 7**). The IIE signature was also predictive for residual cancer burden (RCB class 0 or 1 versus 2 or 3) across all cohorts where annotations were available (**Supplemental Table 8**), with the highest tertile identifying cases with an up to 80% likelihood of RCB 0-1 disease. Although tertiles of the signature identify groups with consistent response rates, we found that response rates are broadly consistent across a wide number of thresholds of IIE signature (**Extended Data** Figure 6). Thus, tailored thresholds can be selected to identify other groups of interest – for example, a rule out threshold can identify 12% of HER2- cases (132 / 1115) across cohorts with a negative predictive value for pCR of 95.5% (representing 6 complete responses out of 132 cases). Explainability analysis demonstrated that the attention scores of the model were highly dependent on tumor cell density (**Figure 5A**).

**Figure 5.**
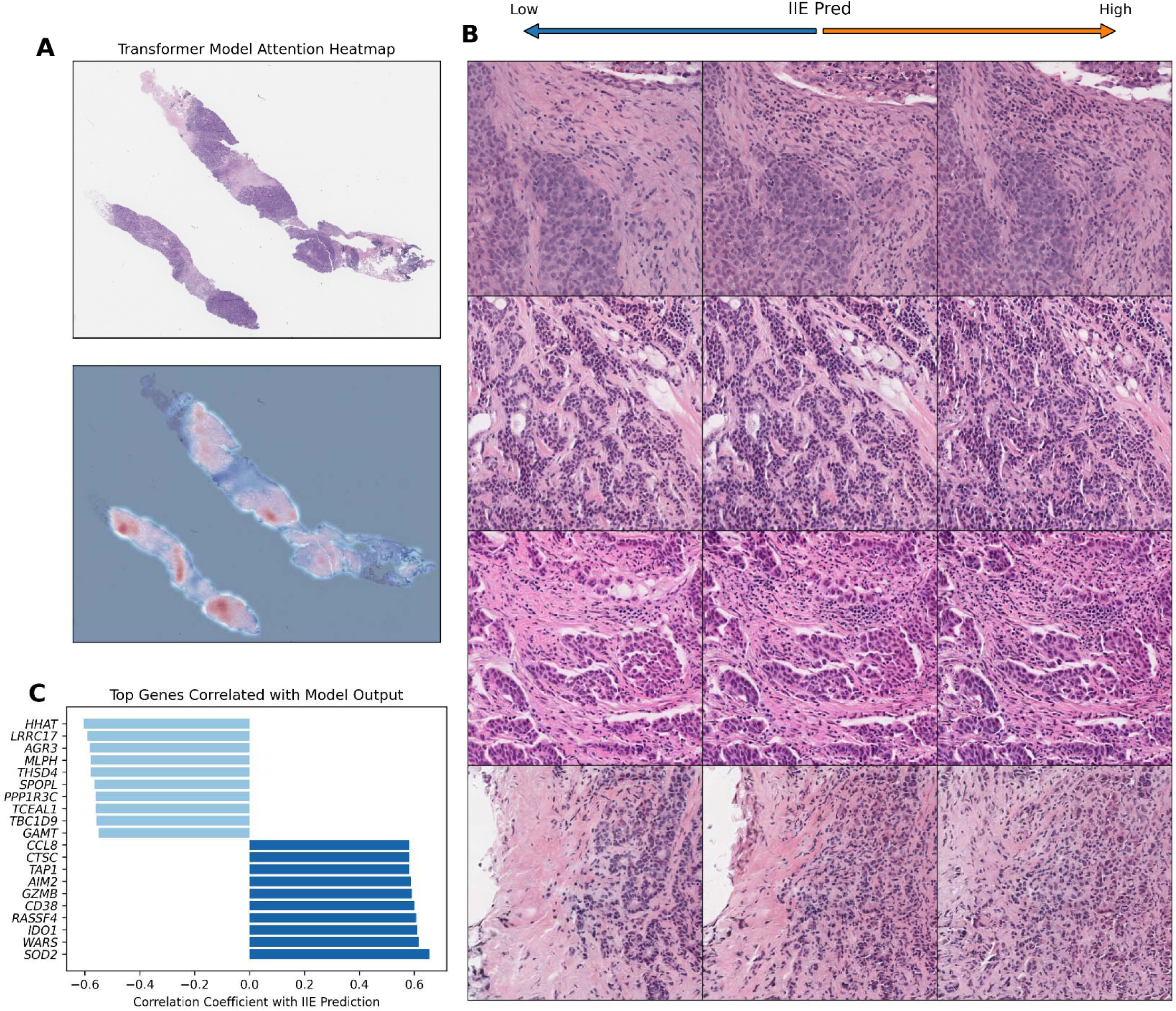
Explainability for IIE Signature Predictions. **A.** An example heatmap of model attention scores demonstrates that attention is highly dependent on tumor cell density. **B.** HistoXGAN was used to generate synthetic histology images from feature vectors, with gradient descent applied to create vectors associated with low / high prediction for the IIE signature. Consistent findings include an increase in tumor infiltrating lymphocytes and less differentiation / tubule formation with high IIE signature prediction. **C.** Genes associated with IIE signature prediction (here calculated in the I-SPY2 dataset) – notably several immune related genes including *CD48* and *GZMB* were associated with a high IIE prediction. **Abbreviations: HistoXGAN**, Histology feature eXplainability Generative Adversarial Network; **I- SPY2**, Investigation of Serial Studies to Predict Your Therapeutic Response with Imaging and Molecular Analysis 2.

Images with higher density of TILs as well as less differentiation resulted in higher prediction for IIE signature (**Figure 5B**). The genes most associated with IIE predictions from histology included a number of immune-related genes such as *CD48* and *GZMB* (**Figure 5C**).

### Gene Expression Signatures for Response to Specific Treatments

We then evaluated the ability of histology derived signatures to predict specific response to immunotherapy in HER2- patients, as treatment with / without immunotherapy was well represented in our training set and validation cohorts. The ideal treatment-specific predictor would have discriminative value in patients receiving a treatment, but no predictive value among patients who do not receive a treatment. When evaluating prediction of response to chemotherapy + immunotherapy versus chemotherapy alone in I-SPY2, the accuracy of prediction appears nonlinear, with some signatures predicting response only in the chemotherapy + immunotherapy arm (**Extended Data** Figure 7A). Interestingly, a signature for the HER2-enriched breast cancer subtype^25^ achieved the highest prediction for immunotherapy + chemotherapy response (AUC 0.638, 95% CI 0.587 - 0.688, p < 0.001) while maintaining no predictive value for chemotherapy response (with AUC < 0.55 for chemotherapy). This signature was derived from HER2-enriched PAM50 subtype (representing a subset of ∼50% of HER2+ cases^26^), and the histology model for this signature does not accurately distinguish HER2 amplification, but instead captures reproducible histologic features that are common in HER2-enriched disease (**Extended Data** Figure 8). This signature was predictive for immunotherapy response in the Yale durvalumab cohort (0.681, 95% CI 0.507 - 0.855, p = 0.025) but not predictive in the other external chemotherapy cohorts. Patients in the lowest tertile for this signature had poor response to immunotherapy regimens (**Supplemental Table 9**, **Extended Data** Figure 7B) – with 30% lower rates of pCR and 25% lower rates of RCB 0-1 responses (**Supplemental Table 10**) compared to higher tertiles in the immunotherapy cohorts. The histologic features leading to a high prediction for this HER2 signature included stromal remodeling and a polymorphic infiltrate of TILs, neutrophils, and spindle / dendritic cells (**Extended Data** Figure 7C).

HER2+ patients represented a minority of patients in our training / validation cohort, and the treatment regimens received were variable, with some cohorts receiving pertuzumab and trastuzumab, and some treated with trastuzumab alone. In I-SPY2, there was a linear relationship between signatures and response prediction in the HER2- and HER2+ cohorts (**Extended Data** Figure 7D); however, a TNFα signature was the only predictive marker for response in the HER2+ group after false discovery correction (AUC 0.668, 95% CI 0.563 - 0.772 p = 0.002; **Supplemental Table 11**). This signature was predictive in the UChicago HER2+ cohort (AUC 0.697, 95% CI 0.544 – 0.850, p = 0.009) and in the CALGB 40601 study (AUC 0.626, 95% CI 0.542 – 0.710, p = 0.003).

Tertiles of this TNFα signature distinguished response, with a pCR rate of over 55% in all cohorts (**Extended Data** Figure 7E) – and over 80% of patients in the highest tertile had RCB 0 or 1 disease (**Supplemental Table 12**) when annotations were available. Predictions for this signature were strongly dependent on the presence of immune infiltrate / TILs (**Extended Data** Figure 7F).

The CALGB 40601 and 40603 studies present an opportunity to test associations with gene expression for other non-standard treatments in breast cancer. In particular, we evaluated specific predictors to bevacizumab and carboplatin response in TNBC, and lapatinib and trastuzumab in HER2+ breast cancer (**Extended Data** Figure 9). Interestingly, we found several claudin-low / mesenchymal subtype signatures^27^ were specifically associated with bevacizumab response in

CALGB 40603, but not in the arms including chemotherapy alone. Additionally, a basal subtype signature^25^ was specifically associated with response to carboplatin. Conversely, in the HER2+ CALGB 40601 study, there was no clear association with a signature and response to either lapatinib or trastuzumab.

### Multiomic Response Prediction from Digital Histology and RNA Expression Data

Although digital histology could be widely utilized for response prediction with minimal added cost, we assessed whether we could combine digital pathology predictions with gene expression to improve response prediction (**Figure 6A**). We trained single-modality RNA-based self-normalizing neural network models separately in HER2-negative (n = 742) and HER2-positive (n = 245) breast cancer cohorts from the I-SPY2 trial, incorporating 17,274 genes where matched expression data was available in both I-SPY2 and CALGB trials. Batch effects were estimated for each gene using a hierarchical Bayesian model fit on the TNBC and CALGB 40603 cohorts, and subsequently applied to the full I-SPY2 microarray data to yield similar gene distributions in the I-SPY2 / CALGB cohorts (**Extended Data** Figure 10). RNA models were optimized with five-fold cross validation in I-SPY2, and subsequently tested in the TNBC CALGB 40603 cohort (n = 234) and HER2+ CALGB 40601 cohort (n = 195). For each subgroup, we additionally integrated patient-level predictions from the IIE signature, and fit a logistic regression with the gene expression model and pathology predictions to integrate these two forms of data for prediction. For the HER2- training / testing cohorts, the combined model (AUROC 0.726 in I-SPY2; 0.730 in CALGB 40603) outperformed pathology alone (AUROC 0.701, p comparison 0.039 in I-SPY2; AUROC 0.640, p comparison 0.013 in CALGB 40603); whereas prediction from RNA expression alone was not significantly better than pathology (AUROC 0.739, p comparison 0.536 in I-SPY2; AUROC 0.705, p comparison 0.318 in CALGB 40603; **Figure 6C; Supplemental Table 13**). A similar trend was seen in the HER2+ training cohort, although differences were not significant (AUROC 0.756 / 0.776 / 0.785 for the path / RNA / combined models); whereas the RNA and combined models both outperformed pathology in the CALGB 40601 validation cohort (AUROC 0.686, p comparison 0.006 for RNA; AUROC 0.637, p comparison 0.008 for combination; **Figure 6D**). SHAP plots of individual gene contributions to predictions demonstrated that immune chemokines (*CXCL13*) and MHC markers (*HLA-DPB2*, *HLA-DQB1*) were among the top 20 markers positively associated with response in HER2- patients (**Figure 6E**), whereas *GRB7* expression (often co-amplified with *ERBB2*) was among the top genes associated with HER2+ response. SHAP plots of gene groups defined by Hallmark gene sets demonstrated that immune gene sets such as the Allograft Rejection and proliferation set such as G2-M Checkpoint and E2F Targets contributed to positive response prediction for both models, whereas estrogen response pathways were negatively associated with response prediction (**Figure 6 G&H**).

**Figure 6.**
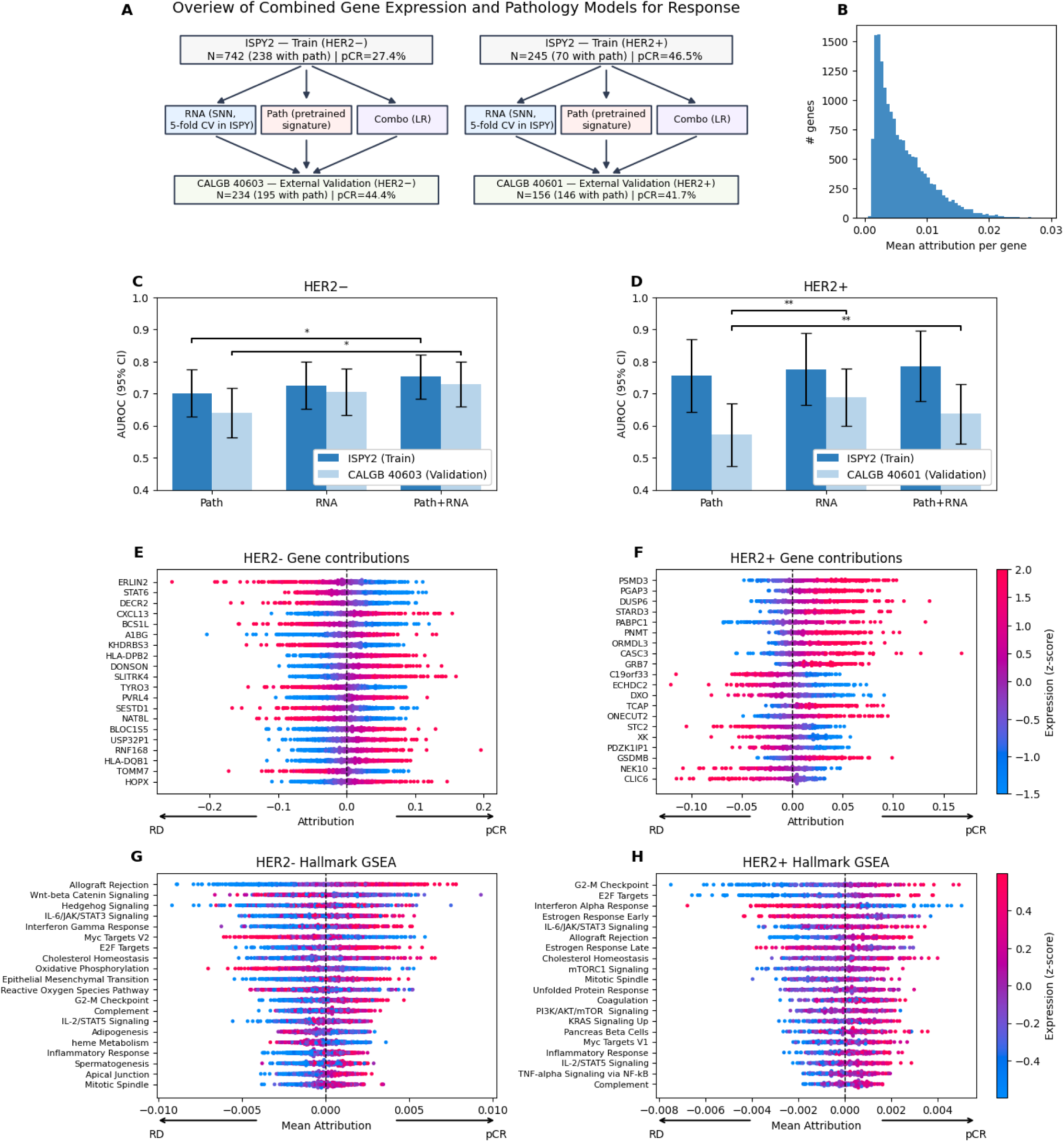
Multiomic Prediction of Pathologic Complete Response from Gene Expression and **Pathology Data. A**, Overview of analysis workflow. Separate RNA-based neural network models were trained in HER2− and HER2+ I-SPY2 cohorts, with 5-fold cross validation used to estimate training performance, and predictions combined with the pre-trained pathology IIE signature prediction via logistic regression to generate multimodal predictors. All three models (RNA, pathology, RNA+pathology) were then validated in CALGB 40603 (HER2−) and CALGB 40601 (HER2+) cohorts. **B**. Distribution of absolute gene attribution magnitudes, calculated via integrated gradients, for all genes for the HER2- model, illustrating that predictions are not dominated by a small subset of genes. **C & D**, Model performance and confidence intervals in I-SPY2 (training) and CALGB (validation) datasets, with significance of model comparisons assessed by DeLong’s test. **E & F**. SHAP plots of top 20 genes by mean integrated gradient attribution in the HER2− and HER2+ cohorts, with each point representing a single patient, with the color indicating standardized expression of the referenced gene, and x-axis indicating contribution direction and magnitude of that gene to predicted pCR probability. **G & H**. SHAP plots of top 20 Hallmark pathways ranked by mean integrated gradient attribution across genes included in the pathway in the HER2− and HER2+ cohorts, colored by average expression score for the pathway. * indicates p < 0.05 ** indicates p < 0.01 **Abbreviations: AUROC**, area under the receiver operating characteristic curve; **I-SPY2**, Investigation of Serial Studies to Predict Your Therapeutic Response with Imaging and Molecular Analysis 2; **CALGB**, Cancer and Leukemia Group B; **CV**, cross validation; **SNN**; self-normalizing neural network; **LR**, logistic regression; **HER2**, human epidermal growth factor receptor 2; **pCR**, pathological complete response; **RD**, residual disease; **GSEA**, gene-set enrichment analysis; **SHAP**, SHapley Additive exPlanations.

## Discussion

This study demonstrates the feasibility and clinical impact of transformer-based predictions of gene expression signatures from standard hematoxylin and eosin (H&E)-stained slides in breast cancer. By focusing on the prediction of biologically validated breast cancer gene expression signatures, rather than individual genes, our study addresses both the interpretability challenge and the frequent difficulty in achieving strong predictive performance at the single-gene level. While prior works have demonstrated moderate success in reconstructing gene expression from H&E images, the use of foundational models for feature extraction and a transformer-based architecture to directly predict expression and signatures appears to better handle the massive size and complexity of whole-slide images. Transformers, by incorporating attention mechanisms, allow the model to selectively emphasize tiles of histology images that provide the most relevant information regarding gene expression patterns^28^. This enhanced ability to learn spatially informed representations likely underpins the superior performance of TIGER for gene expression prediction and could enable further histology-driven gene expression biomarkers across a spectrum of cancers. Furthermore, pretraining models to predict clinically relevant breast cancer gene signatures allows for the reduction of digital histology images into a small set of biologically meaningful variables, which can be readily associated with outcomes of interest. Indeed, we found that this approach performs as well or better than models trained to directly predict response in the I-SPY2 cohort and can be used to discover treatment-specific biomarkers even when digital imaging data may be too sparse to train models from scratch. Previous studies have clarified the additive value of DNA mutational status, clinical variables, and digital histology TIL analysis to RNA expression data – finding that no data significantly added to RNA expression^29^. In contrast, we have demonstrated that digital histology, using modern foundational models and pretrained to predict gene expression, performs similarly to a deep learning RNA expression model, and can provide additive value in response prediction to RNA alone – at least in the case of HER2- disease.

The most well-recognized digital image-based marker of response in breast cancer is the quantification of tumor-infiltrating lymphocytes (TILs), which has been linked to pathologic complete response to chemotherapy^30^, immunotherapy^31^, and even benefit of endocrine therapy^32^. However, despite numerous attempts to standardize assessment of TILs, there is significant variability of TIL quantification between pathologists, limiting real world application^33,34^. There are ongoing efforts to replicate manual TIL quantification that achieve similar predictive accuracy as pathologist assessment^35^ - but gene expression signatures may outperform TILs in response / outcome prediction in breast cancer^36,37^. Similarly, computational approaches may identify the precise local / global patterns of immune cell distribution or even phenotypes of individual immune cells^38^ that are predictive of response. The fact that the most predictive signatures in this analysis were largely immune signatures adds to the biologic plausibility of reconstructed signatures for response prediction.

Moreover, we found that the IIE signature—encompassing proliferative, apoptotic, and interferon-related gene features derived from ER+ cancers—was a robust predictor of response across multiple cohorts. This signature has demonstrated strong correlation with TILs in prior studies^37^, and was associated with TIL infiltrate in our explainability analysis. Similarly, a TNFα signature largely based on immune infiltrate was predictive of response in HER2+ patients. Markers of grade / proliferation were also associated with higher prediction of the IIE signature, and IIE prediction was also associated with hormone receptor negativity. Although it has been well recognized that immune infiltrate, grade, and hormone receptor status are associated with response to therapy – this signature may capture the precise combination of these features that is optimally predictive. Indeed, this signature was more predictive than clinical variables or TILs in our analysis. The clinical utility of this approach appears greater than other approaches using AI-derived TILs alone. For example, an analysis of SWOG S0800 of mixed HR+/HR- HER2- breast cancer found a 20% difference in pCR rate between high / low AI TILs; whereas here we found a >45% difference in pCR rate in the low / high tertiles of IIE signature in the mixed HR+/HR- I-SPY2 and UChicago cohorts. Furthermore, we demonstrate that results for these predictions are reliable across multiple slides from the same tumor, and even between FFPE and frozen specimens – overcoming one of the largest barriers to clinical implementation of TIL assessment for response prediction. Of note, there are clinical trials that are evaluating de-escalated treatments for early stage TNBC based on TIL assessment – including the ETNA trial (NCT6078384) evaluating no adjuvant therapy in patients with high TILs, and the OPTImaL trial (BOOG 2024-03) evaluating chemotherapy versus observation in a similar cohort - and this approach could perhaps serve as a more reproducible surrogate for such trials.

An additional key contribution of this work is the demonstration that specific histology-derived signatures can differentiate responses to immunotherapy versus chemotherapy alone. Although PD- L1 status can predict response to such therapy in the metastatic setting, it has failed as a predictive biomarker in both triple-negative and HR+ early stage breast cancer^39,40^. Conversely - the DetermaIO^41^ and ImPrint^42^ signatures have shown promise in prediction of response specifically in patients receiving immunotherapy. Intriguingly, our results suggest that a HER2-associated signature was predictive of immunotherapy benefit in HER2- cohorts of I-SPY2 and a small external cohort, but not predictive of chemotherapy response alone in any cohort. Predictions for the HER2 signature were correlated with predictions for several immune signatures^43,44^, but remained very distinct from the IIE / TNFα signatures – perhaps suggesting that this histologic signature is identifying a unique immune element that is essential for immunotherapy response. The DetermaIO signature was included in our 775 signatures reconstructed from histology – and was moderately correlated with the HER2 signature selected for IO response prediction. However, the histology-derived version of the DetermaIO signature – unlike our selected HER2 signature – was associated with response in all chemotherapy cohorts in this study. The prediction of this signature from histology was moderate-strong (correlation coefficient of 0.64) – so these findings may not reflect the performance of DetermaIO assessed through true gene expression profiling – but it will be important to see if DetermaIO remains predictive in larger populations.

These findings suggest that gene expression signatures derived from histology can provide a biologically anchored framework to identify treatment-specific biomarkers, particularly in the context of non-standard therapies. We found that several claudin-low signatures were selectively associated with response to bevacizumab but not to chemotherapy alone in the CALGB 40603 TNBC cohort.

This raises the possibility that claudin-low may reflect a vascular phenotype particularly susceptible to VEGF inhibition – and studies have previously postulated that claudin-low cells can promote tumor vascular permeability and may respond favorably to anti-vascular agents^45,46^. Additionally, a preclinical model demonstrated that bevacizumab can upregulate claudin expression through modulation of tumor vasculature and intercellular junctions, suggesting a mechanistic link that could explain claudin-low histologic signatures as biomarkers for anti-angiogenic therapy^47^. Previous analysis had found that a breast cancer lung-metastasis signature best predicted bevacizumab response in CALGB 40603^37,48^ – this signature was strongly correlated (Pearson r 0.78) with one of the predictive claudin signatures we identified. Additionally, a basal-like signature was specifically associated with response to carboplatin, consistent with platinum sensitivity observed in TNBC, and could be used as a biomarker to select additional patients such as ER-low cases for carboplatin – and unlike prior histology HRD biomarkers^49^, this represents a specific signal of carboplatin sensitivity rather than general treatment sensitivity. Collectively, these results highlight the promise of leveraging expression signatures inferred from routine histology to identify biomarkers and better understand the biology underlying response to specific treatments.

Despite the promising results, several limitations warrant discussion. First, our study relied on heterogeneous retrospective datasets – and it is possible that our assessments will not fully generalize in other settings. We did find general consistency in prediction across the tissue specimen types and slide scanners included in this study, but consistency and accuracy for response prediction was lower in frozen specimens. However, many older studies have only digital images of frozen specimens available for analysis, and it is reassuring that our model provides moderate to highly consistent predictions from FFPE and frozen specimens from the same patient. Although this study included nearly 500 immunotherapy treated patients, there were limited external cohorts of such patients available for validation of immunotherapy-specific response prediction. Similarly, our training and validation cohorts of HER2+ breast cancer were less robust, and the discriminative value of the TNFα signature was not as high as was seen for the IIE signature. Nonetheless, we believe this approach can serve as a foundation for future studies, where histology-derived gene expression can rapidly generate a suite of biomarkers across retrospective studies – for both hypothesis generation and personalization of treatment. Finally, although we found our pathology signatures outperformed predictions from standard clinical biomarkers, TIL assessment and quantitative assessment of ER, PR, Ki-67, and HER2 FISH testing were either not available or only available in small cohorts.

Machine learning studies have accurately predicted response from such markers^50^ – and it will be important to assess if our digital histology approach has additive value to standard quantitative biomarkers. TIL assessment across this large cohort of > 4,000 digital images is ongoing to enable further comparison.

In conclusion, this study provides evidence that transformer-based histology analysis can accurately predict a range of biologically and clinically relevant gene expression programs in breast cancer. By capturing key signals related to proliferation, hormone receptor status, immune infiltration, and more, TIGER-derived signatures may help guide neoadjuvant treatment decisions and identify patients most likely to benefit from immunotherapy. With ongoing improvements in deep learning architectures, annotation tools, and interpretability methods, digital pathology is poised to accelerate the realization of more precise, cost-effective, and readily scalable molecular diagnostics in oncology.

## Materials and Methods

### Data Sources and Image Extraction

Breast cancer cases from TCGA were used for training and cross validation of gene expression and signature prediction models. Slides and associated clinical data were accessed through the Genomic Data Commons Portal (https://portal.gdc.cancer.gov/). Neoadjuvant datasets used for response prediction include cases from the I-SPY2 clinical trial (NCT01042379, data from which can be obtained through an approved data request submitted to i-), a single arm phase II trial of durvalumab + neoadjuvant chemotherapy from Yale Cancer Center (NCT02489448), patients from the CALGB 40601 (NCT00770809) and 40603 (NCT00861705) clinical trials, and prospectively consented patients receiving standard of care neoadjuvant chemotherapy at University of Chicago from 2006 - 2021. This analysis was reviewed by the University of Chicago IRB under approved protocol 22-0707.

Samples in the above cohorts with image tiles extractable with our SlideFlow pipeline were included in the analysis, with a CONSORT diagram of exclusions per cohort illustrated in **Extended Data** Figure 1. Slide images were extracted using the Slideflow pipeline with a tile size of 224px per 224µm to achieve an effective optical resolution of 10X, with filtering to remove tiles with > 60% grayspace and additional Otsu thresholding and Gaussian blur based filtering to remove out of focus tiles / pen marks as illustrated in **Extended Data** Figure 251. No tumor region of interest annotations were performed in this analysis.

### Genomic Feature and Signature Prediction

FPKM upper quartile normalized mRNA expression data was obtained as previously published from TCGA and CPTAC (https://portal.gdc.cancer.gov/); Salmon processed upper-quartile normalized RNA expression data was obtained from CALGB 40601 (GEO GSE116335) and CALGB 40603 (GEO GSE154524) and normalized microarray based mRNA expression was obtained from I-SPY2 (GEO GSE194040). All gene expression values were included (analysis was not limited to protein coding gene expression). Gene expression values were log-transformed to stabilize variance and approximate a normal distribution prior to downstream statistical analysis. Specifically, values were transformed as log_g:_rx + 1) to accommodate zero counts. To enable consistent gene annotation across datasets, gene identifiers were harmonized using the GENCODE v22 reference annotation and the Python package mygene^52^. ENSEMBL gene IDs were mapped to HGNC-approved gene symbols and Entrez Gene IDs using the mygene query service. We also utilized gene expression signatures (n = 775) previously computed in the TCGA, CALGB 40601, and 40603 datasets that distill gene expression into biologically relevant patterns that comprehensively characterize breast cancer biology and are more prognostic than individual gene expression^36,37,53^. Previously published version of these signatures are used to ensure consistency with prior analyses. Clustering of these signatures into subgroups was performed via HDBSCAN with a correlation distance metric, a minimum of 10 samples, a minimum cluster size of 20.

To improve prediction of gene expression signature prediction in breast cancer, we assessed a number of histology-specific foundational models (CTransPath^10^, HistoSSL^54^, and UNI^11^) and backbones for feature aggregation (attention-multiple instance learning^12^, transformer-based multiple instance learning^14^, and the Bistro transformer architecture^14^) all with a multilayer perceptron head and MSE loss, to develop an optimized approach for expression prediction – in a pipeline we term transformer-based hIstology-driven Gene Expression Regressor (TIGER). The optimal feature aggregation architecture was adapted from a prior transformer-based framework^14^, in which each instance is first projected into a higher-dimensional embedding space via a learned linear transformation. A learnable class token is prepended to the sequence, with the core of the model comprised of a stack of transformer blocks, each consisting of multi-head self-attention and a feed- forward network, with class-token pooling used for prediction. Attention computations are chunked to improve memory-efficiency, enabling inference on large slides images for the large number of genes predicted in this analysis.

There exist several published state of the art methods for predicting bulk gene expression (as opposed to signatures). These include DeepPT^8^ – which consists of an autoencoder followed by multilayer perceptron and SEQUOIA^9^ which applies K-means clustering followed by a linearized transformer. We compared TIGER to these two alternative architectures for the task of bulk gene expression prediction, using UNI features as the basis for all three models, with five fold cross validation in TCGA computed for 60,660 genes (although predictive accuracy only assessed in genes with non-zero variance), and independent prediction from the TCGA trained models CPTAC, as well as for 27,851 matching genes in the CALGB 40601 / 40603 cohorts, and in the I-SPY2 cohort for expression of 15,979 matching genes on the Agilent microarray platform. For individual gene expression prediction, models were trained for 200 epochs at a learning rate of 0.001. For signature prediction, the optimal performance in TCGA was seen at epoch 8 at learning rate of 0.001 on cross validation, so a full model for external testing was trained using these parameters across the full dataset. For consistency of signature predictions, the two-way mixed effects intraclass correlation coefficient i.e. ICC(3,k) was calculated in the I-SPY2 dataset – in which patients had multiple samples – including some with a mix of both FFPE and frozen tissue.

### Application of Signature Predictions to External Cohorts

A fixed, optimized TIGER model for prediction of 775 signatures was trained in the TCGA-BRCA cohort. To limit statistical testing for associations with outcome, signatures that were predictable with a correlation coefficient of < 0.5 from initial five-fold cross validation in TCGA, or that were highly correlated with other signatures (correlation coefficient > 0.95 when calculated across all patients in the I-SPY2 dataset) were discarded from the analysis – resulting in a set of 61 signatures (**Extended Data** Figure 3, **Supplemental Table 5**). The AUROC of each signature for prediction to pathologic complete response was assessed both overall and in relevant patient subgroups – including those treated with chemotherapy versus chemotherapy + immunotherapy, and those treated with or without HER2-targeted therapy. Optimal signatures for 1) response to neoadjuvant chemotherapy in HER2- patients, 2) response to neoadjuvant therapy in HER2+ patients receiving trastuzumab, and 3) ***specific*** prediction of response to immunotherapy selected in the I-SPY2 cohort. The first two signatures were chosen based on the highest AUROC in the I-SPY2 cohort; for immunotherapy-specific response prediction we selected the signature with highest AUROC in the chemotherapy + immunotherapy group with low predictive value (AUROC between 0.45 and 0.55) in the chemotherapy group. To select a threshold applicable to clinical practice, we identified thresholds based on tertiles of predictions in I-SPY2. Accuracy of predictions were compared to a logistic regression from clinical factors – including age, tumor stage, nodal stage, grade, HR status, and HER2 status – fit on the I-SPY2 dataset. Additionally, predictions were compared to pathologist TIL annotations in cohorts where this data was previously calculated, and TIL annotations were annotated on the UChicago cohort by a study pathologist (G.K.)^55^.

### Multiomic Response Prediction

Gene expression microarray data from the TNBC subset of the I-SPY2 was normalized against the TNBC CALGB 40603 study. Batch effects were estimated for each gene using the ComBat^56^ algorithm as implemented in the neuroHarmonize package^57^. This correction was then applied to the full I-SPY2 dataset. For response prediction we implemented a self-normalizing neural network (SNN) for omics data, consisting of four sequential fully connected layers with 256 units each, ELU activation, and alpha dropout for regularization, and unit-normalized gene expression data of common genes between these datasets was used as model input. Models were trained with 5-fold cross validation in TCGA, with the optimal number of epochs selected for external testing based on average AUROC for response on cross validation. For multimodal analysis, we combined RNA model predictions with the IIE signature predictions from histology, using a logistic regression model fit without regularization or tuning. Final performance was assessed in the held-out external validation CALGB 40601 / 40603 studies.

### Explainability of Predictions

Heatmaps of attention were generated across whole slide images to identify areas of slides that contributed most to model predictions. Additionally, we applied our HistoXGAN^15^ explainability architecture – which generates representative pathology images from foundational model features. Gradient descent was used to modify the foundational model feature vector to create representative images of high / low signature prediction. Histology images were reviewed by two breast cancer pathologists to identify the histologic features representative of low / high predictions. Finally, the association of predictions of signatures with individual genes in the I-SPY2 dataset was assessed.

For explainability analysis of RNA models, feature contributions to model predictions were quantified using integrated gradients^58^ as implemented in the Captum package^59^. For each sample, attributions were computed relative to a zero vector baseline and averaged across the I-SPY2 training set within each HER2 subgroup. Absolute attribution magnitudes were used to rank features for the global importance distribution. Hallmark gene set scores were computed from MSigDB^60^ by averaging integrated gradient-weighted expression across all genes in a pathway.

### Statistical Analysis

Correlation of true versus predicted gene expression (both individual gene level and signature) was assessed using Pearson correlation coefficient averaged in the held out test set with fivefold cross validation in TCGA, and independently in the CPTAC, CALGB 40601, CALGB 40603, and I- SPY2 cohorts. A t-test was used to compare the average correlation for the top 1000 genes in these two datasets across methodologies.

To obtain an overall estimate of classifier performance across multiple treatment arms, we computed a weighted average AUROC for all aggregates reported in the I-SPY2, CALGB 40601, and CALGB 40603 trials. For each treatment arm, we extracted the AUROC (A_l,_), its estimated variance (l_l,_), and the number of evaluable patients (n_l,_).

The overall weighted AUROC A was computed as:

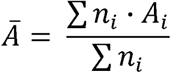

and the corresponding weighted variance of the AUROC as:

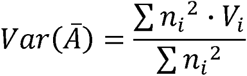

Confidence intervals for AUROC predictions and significance testing (versus a null of 0.50) is computed via normal proximation as per the methodology of Hanley and McNeil^61^. Comparison between predictions from two approaches (signature vs clinical features; signature vs TILs) was assessed with the Delong methodology^62^. Significance for associations in I-SPY2 were adjusted using the Benjamin-Hochberg method^63^ for all primary analyses. All statistical testing was two sided using the alpha = 0.05 confidence level. Analyses were performed in duplicate (representing a technical replicate) with identical results.

## Acknowledgements

Data from Cancer and Leukemia Group B (CALGB) studies 40601 and 40603 were obtained from the Alliance for Clinical Trials in Oncology, a National Clinical Trials Network cooperative group, under Alliance data sharing agreement A152001; those studies were supported in part by the National Cancer Institute of the National Institutes of Health (NIH) under Award Numbers U10CA180821 and U10CA180882 (to the Alliance for Clinical Trials in Oncology). The current study was supported in part by the National Cancer Institute of the National Institutes of Health (NIH) under Award Numbers UG1CA233327, UG1CA189847, UG1CA233329, UG1CA233337, UG1CA233373, UG1CA233180, UG1CA233290, UG1CA233331, and UG1CA233339. Also supported in part by funds from GlaxoSmithKline/Novartis (CALGB 40601) and Genentech (CALGB 40603). Through acceptance of this federal funding, NIH has been given a right to make the Author Accepted

Manuscript publicly available in PubMed Central upon the Official Date of Publication, as defined by NIH. The content is solely the responsibility of the authors and does not necessarily represent the official views of the National Institutes of Health.

This work was also supported by the following research grants: National Cancer Institutes grant K08CA283261 (FMH) Cancer Research Foundation grant (FMH)

Lynn Sage Breast Cancer Foundation (FMH)

Alliance Foundation Trials Special Projects Award (FMH)

National Institute of Dental and Craniofacial Research grant R56DE030958, (ATP) National Cancer Institutes grant R01CA276652 (ATP)

European Commission Horizon grant 2021-SC1-BHC, (ATP) Adenoid Cystic Carcinoma Research Foundation grant (ATP) Cancer Research Foundation grant (ATP)

American Cancer Society grant (ATP)

Department of Defense grant BC211095P1 (FMH, ATP) National Cancer Institutes grant P50CA058223 (CMP) Breast Cancer Research Foundation BCRF-23-127, (CMP) Stand Up To Cancer grant (ATP)

National Cancer Institutes grant P01CA210961 (LJE)

## Data Availability

Data from TCGA including digital histology and clinical annotations used are available from https://portal.gdc.cancer.gov/ and https://cbioportal.org. Data from the I-SPY2, CALGB 40601 / 40603, and Yale clinical trials can be obtained through approved data requests from the respective trial review committees. Data from the UChicago cohort can be obtained via reasonable request to study authors. All code and trained models used for this analysis are available at https://github.com/fmhoward/TIGER.

---

**Extended Data Figure 1.**
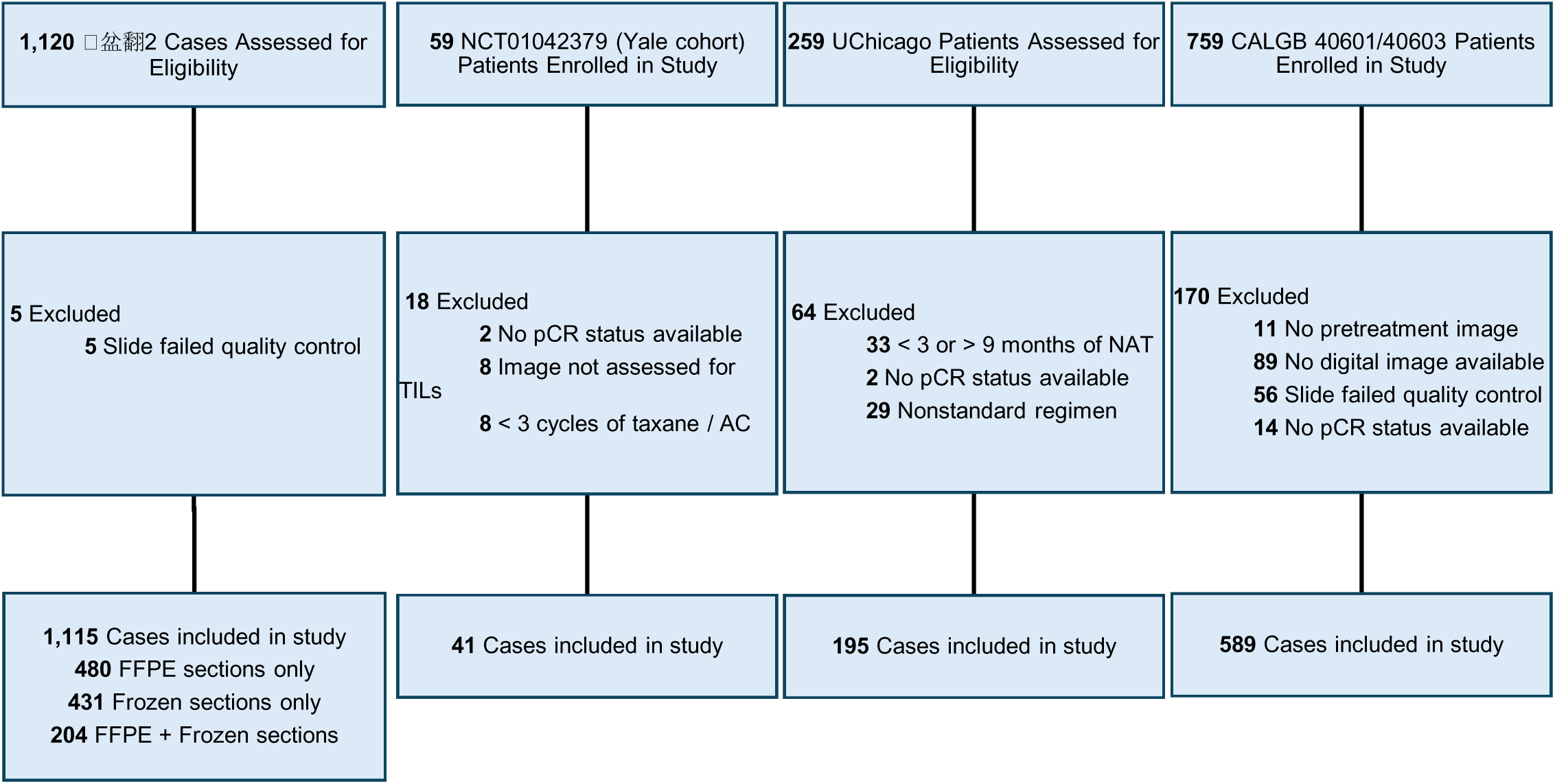
**CONSORT Diagram for Included Patients in Neoadjuvant Setting Abbreviations: I-SPY**, Investigation of Serial Studies to Predict Your Therapeutic Response with Imaging and Molecular Analysis**; CALGB**, Cancer and Leukemia Group B; **pCR**, pathologic complete response; **TIL**, tumor infiltrating lymphocyte; **AC**, doxorubicin and cyclophosphamide; **NAT**, neoadjuvant therapy; **FFPE**, formalin fixed paraffin embedded.

**Extended Data Figure 2.**
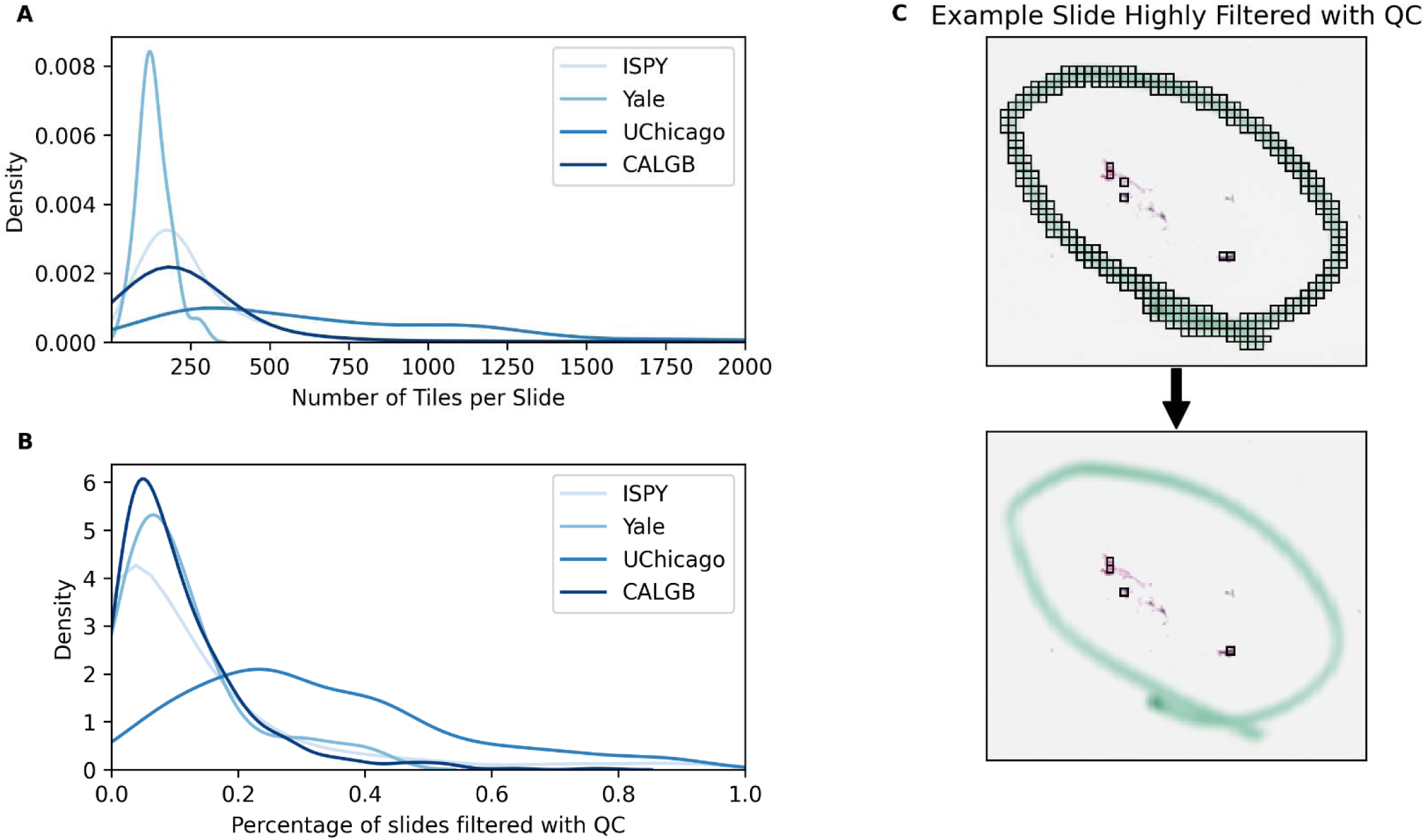
Quality Control and Reproducibility of Signature Predictions. **Otsu** thresholding and Gaussian blur-based filtering was used to perform automated quality control on all included datasets. The number of extracted tiles per slide image (**A**) and the percentage of tiles filtered with quality control (**B**) is shown across each included dataset. Of note, the University of Chicago standard of care specimens generally included multiple biopsy cores per slide image yielding a higher number of tiles per slide image. **C.** The slide image in the I-SPY2 dataset with the highest proportion of tiles filtered with quality control is shown. **Abbreviations: I-SPY**, Investigation of Serial Studies to Predict Your Therapeutic Response with Imaging and Molecular Analysis; **CALGB**, Cancer and Leukemia Group B; **QC**, quality control.

**Extended Data Figure 3.**
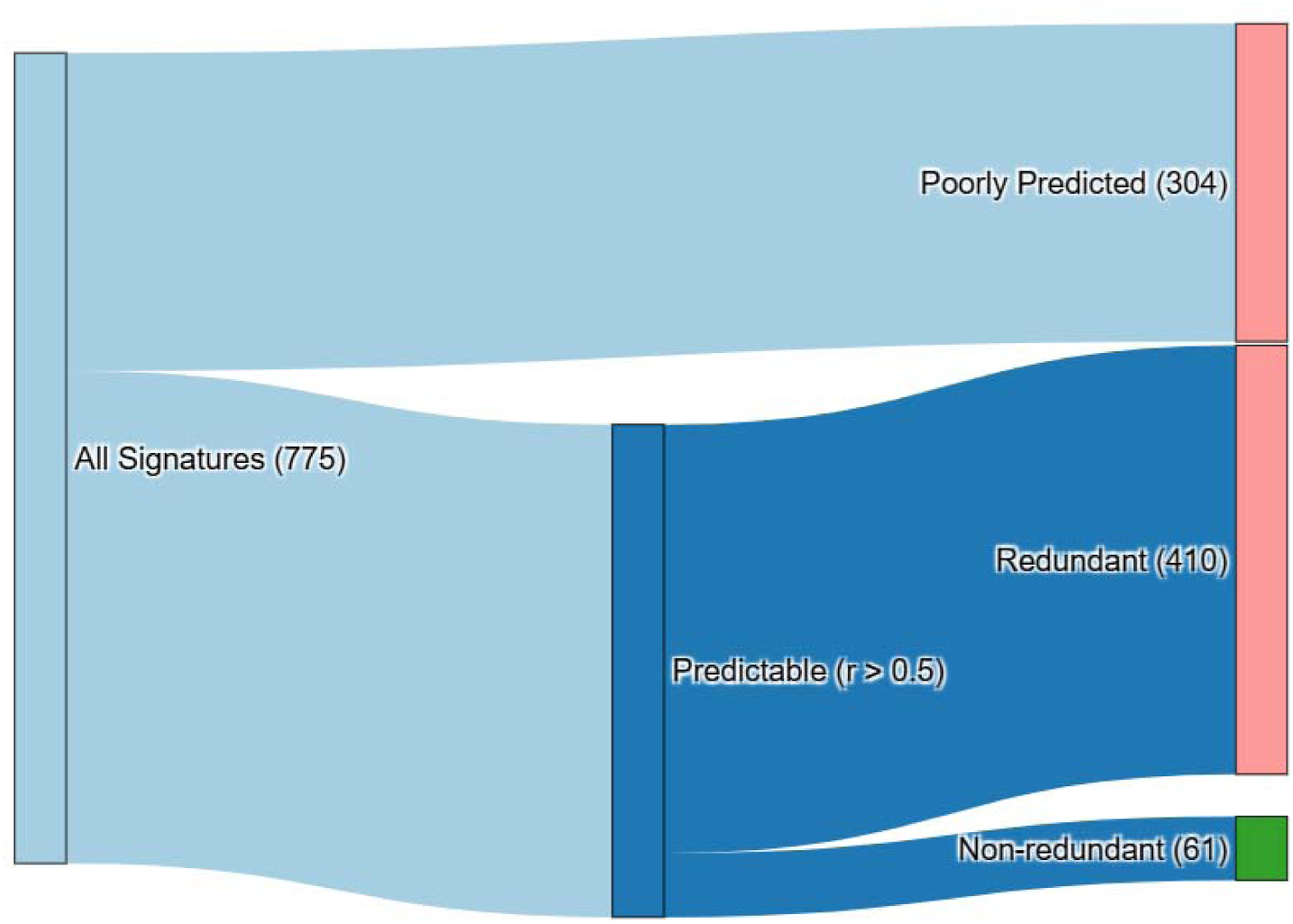
Filtering Pipeline for Gene Expression Signatures. From the original 775 gene expression signatures predicted by our model, 304 signatures are removed from downstream analysis as they were poorly predicted on initial testing (Pearson r < 0.5 with 5-fold cross validation on TCGA). Subsequently, a correlation matrix was constructed from signature predictions across the full I-SPY2 analysis set, and 410 signatures with co-correlation > 0.95 with other signatures were removed, resulting in 61 signatures for downstream analysis. **Abbreviations: I-SPY**, Investigation of Serial Studies to Predict Your Therapeutic Response with Imaging and Molecular Analysis; **TCGA**, The Cancer Genome Atlas.

**Extended Data Figure 4.**
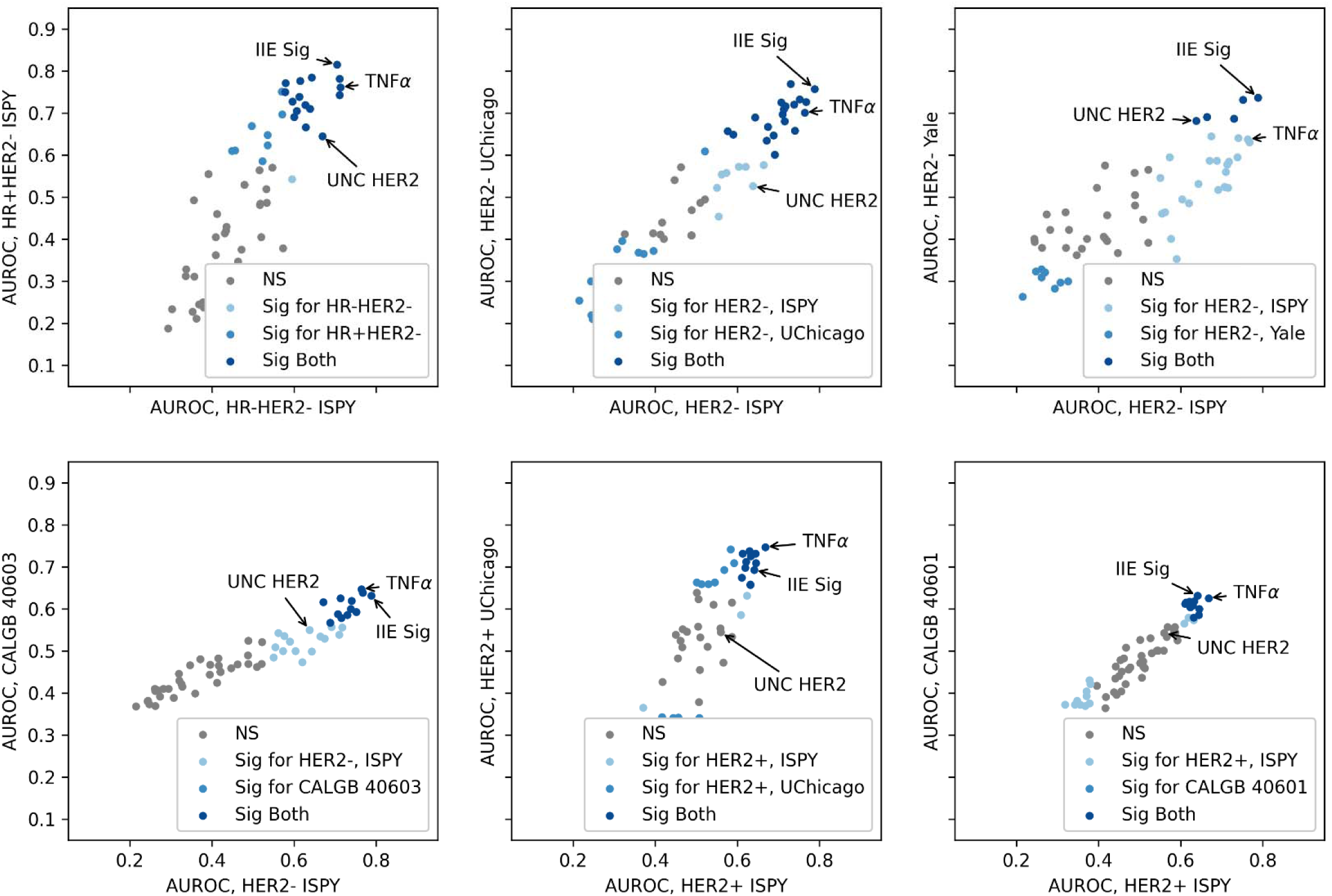
Consistency of Predictive Performance of Histology-Inferred Gene **Expression Signatures Across Similarly Treated Cohorts.** Each plot compares the AUROCs for pathologic complete response prediction using 61 distinct and predictable gene expression signatures inferred from histology, across pairs of independent breast cancer cohorts, including HER2- and HER2+ disease. Signatures demonstrate similar predictive performance across similarly treated cohorts. The IIE signature demonstrates robust accuracy across HER2- cohorts, whereas the UNC HER2 signature is only predictive in immunotherapy treated HER2- cohorts. As these assessments are considered exploratory, false discovery correction was not performed. **Abbreviations**: **I-SPY**, Investigation of Serial Studies to Predict Your Therapeutic Response with Imaging and Molecular Analysis; **CALGB**, Cancer and Leukemia Group B; **AUROC**, area under the receiver operating characteristic curve; **HER2**, human epidermal growth factor receptor 2; **HR**, hormone receptor; **NS**, not significant; **pCR**, pathological complete response; **UNC**, University of North Carolina. **TNF**α, tumor necrosis factor alpha.

**Extended Data Figure 5.**
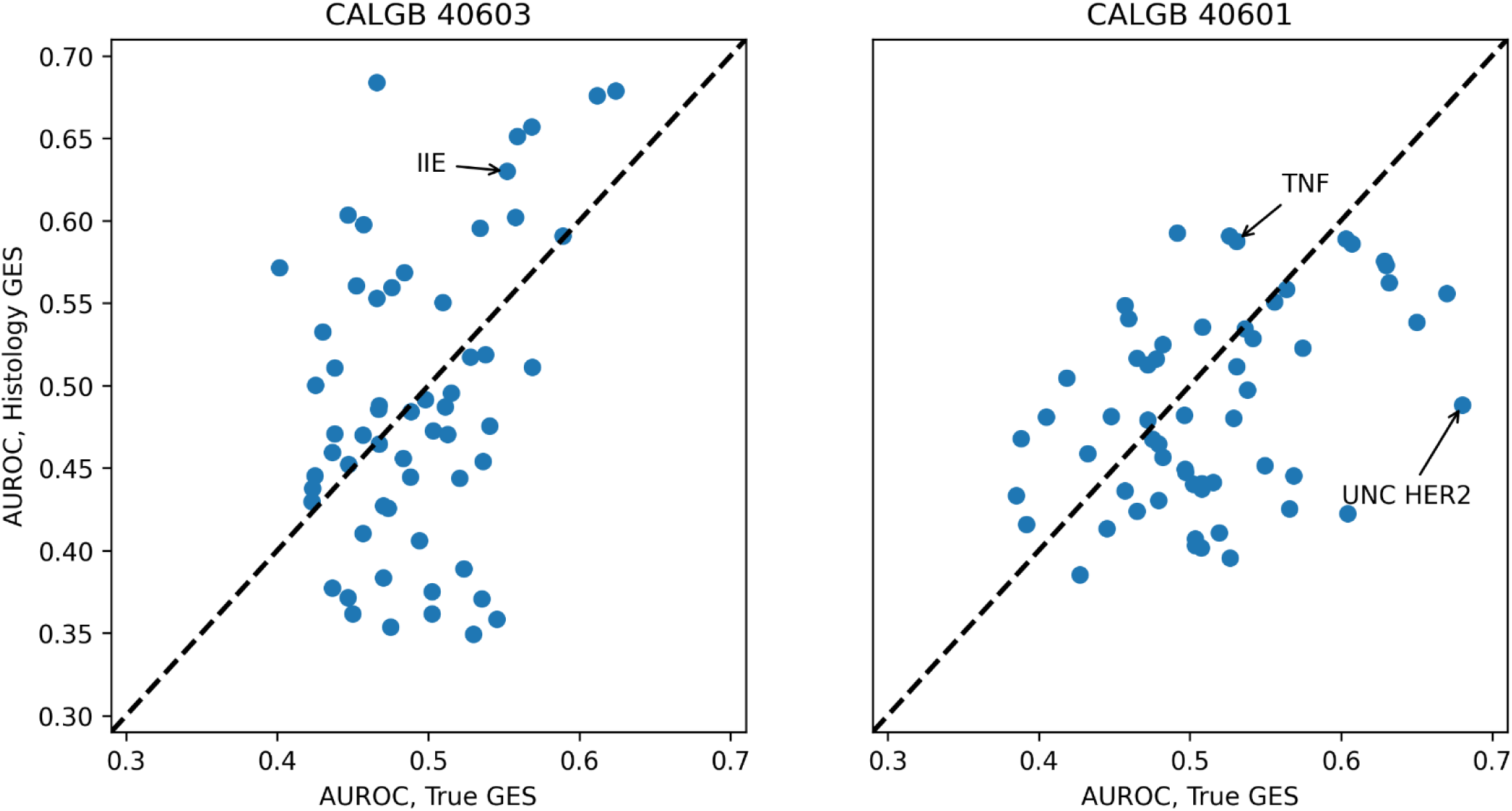
Performance of Histology-Based Prediction of Gene Expression Signatures and Their Predictive Utility for Therapy Response. Scatter plots comparing the AUROC of response prediction for pCR using true vs. histology-predicted gene expression signatures in CALGB 40601 and CALGB 40603. Each point represents the performance of a separate signature – among the 61 highly predictable distinct signatures. The correlation between AUROCs from predicted vs. true signatures was 0.49 in CALGB 40601 and 0.43 in CALGB 40603, demonstrating preserved predictive ranking of signatures across data modalities. Of note, in the HER2+ CALGB 40601 cohort, the UNC HER2 signature was highly predictive from gene expression, but less predictive from digital histology, suggesting the histology version of this signature does not accurately capture differential levels of HER2 expression. **CALGB**, Cancer and Leukemia Group B; **AUROC**, area under the receiver operating characteristic curve; **HER2**, human epidermal growth factor receptor 2; **GES**, gene expression signature; **pCR**, pathological complete response; **UNC**, University of North Carolina. **TNF**α, tumor necrosis factor alpha.

**Extended Data Figure 6.**
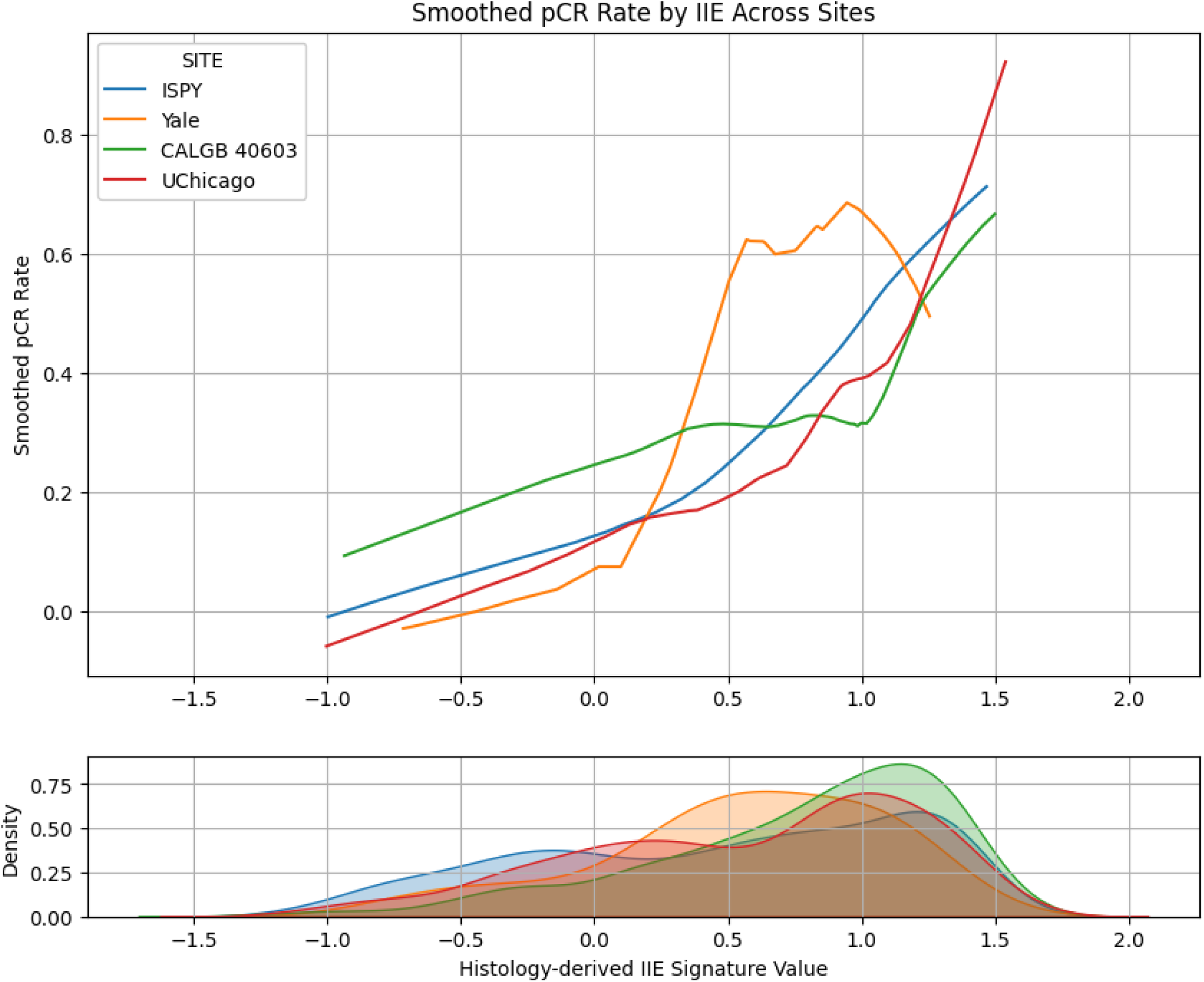
Consistency of Pathologic Complete Response Rate as a Function of **Histology Derived IIE Signature.** Pathologic complete response (pCR) rates were smoothed using locally weighted regression (LOWESS) across percentiles of predicted IIE probability scores derived from FFPE stained digital histology in 1,115 HER2-negative patients across four study cohorts (I- SPY2, UChicago, Yale, CALGB 40603). The top panel shows the smoothed relationship between histology-derived IIE signature and observed pCR rate, demonstrating consistent increases in pCR rates as a function of IIE signature values. The bottom panel displays the kernel density estimate of predicted IIE scores within each cohort, illustrating the distribution of predictions across sites. **I-SPY**, Investigation of Serial Studies to Predict Your Therapeutic Response with Imaging and Molecular Analysis; **CALGB**, Cancer and Leukemia Group B; **pCR**, pathological complete response.

**Extended Data Figure 7.**
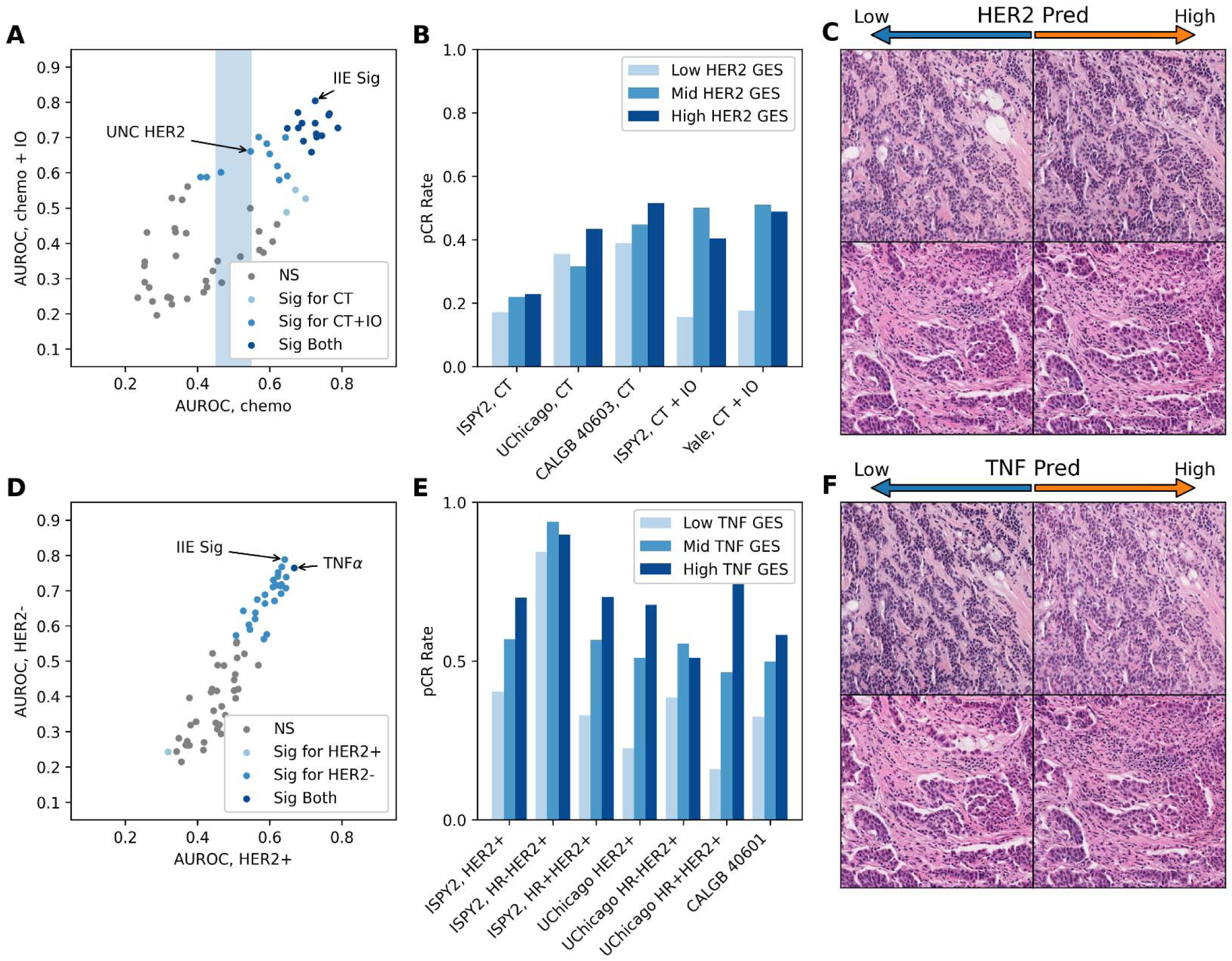
Specific Prediction of Response in Patients Receiving **Immunotherapy and HER2-Targeted Therapy. A.** Some signature predictions demonstrated differential association with response to chemotherapy and response to chemotherapy + immunotherapy in the I-SPY2 cohort. Predictions for a HER2 signature had the highest association with pCR (measured by AUROC) across chemotherapy + immunotherapy arms, while maintaining no association with response to chemotherapy alone. **B**. Pathologic complete response rates in subgroups defined by tertiles of HER2 signature prediction in chemotherapy alone and chemo herapy + immunotherapy cohorts. **C.** Pathologic features associated with HER2 signature prediction included stromal remodeling and a polymorphic infiltrate of TILs, neutrophils, and spindle / dendritic cells. **D.** The association of signatures with response in HER2- versus HER2+ cases was highly linear. Only predictions for a TNFα signature was significantly associated with response in HER2+ patients in the I-SPY2 cohort. **E.** Pathologic complete response rates in subgroups defined by tertiles of TNFα signature prediction in HER2+ cohorts and in subgroups defined by receptor status. **F.** More prominent immune infiltrates were seen in generated images with high TNFα signature prediction. **I-SPY**, Investigation of Serial Studies to Predict Your Therapeutic Response with Imaging and Molecular Analysis; **CALGB**, Cancer and Leukemia Group B; **CT**, chemotherapy; **AUROC**, area under the receiver operating characteristic curve; **HER2**, human epidermal growth factor receptor 2; **HR**, hormone receptor; **IO**, immunotherapy; **GES**, gene expression signature; **pCR**, pathological complete response; **UNC**, University of North Carolina. **TNF**α, tumor necrosis factor alpha.

**Extended Data Figure 8.**
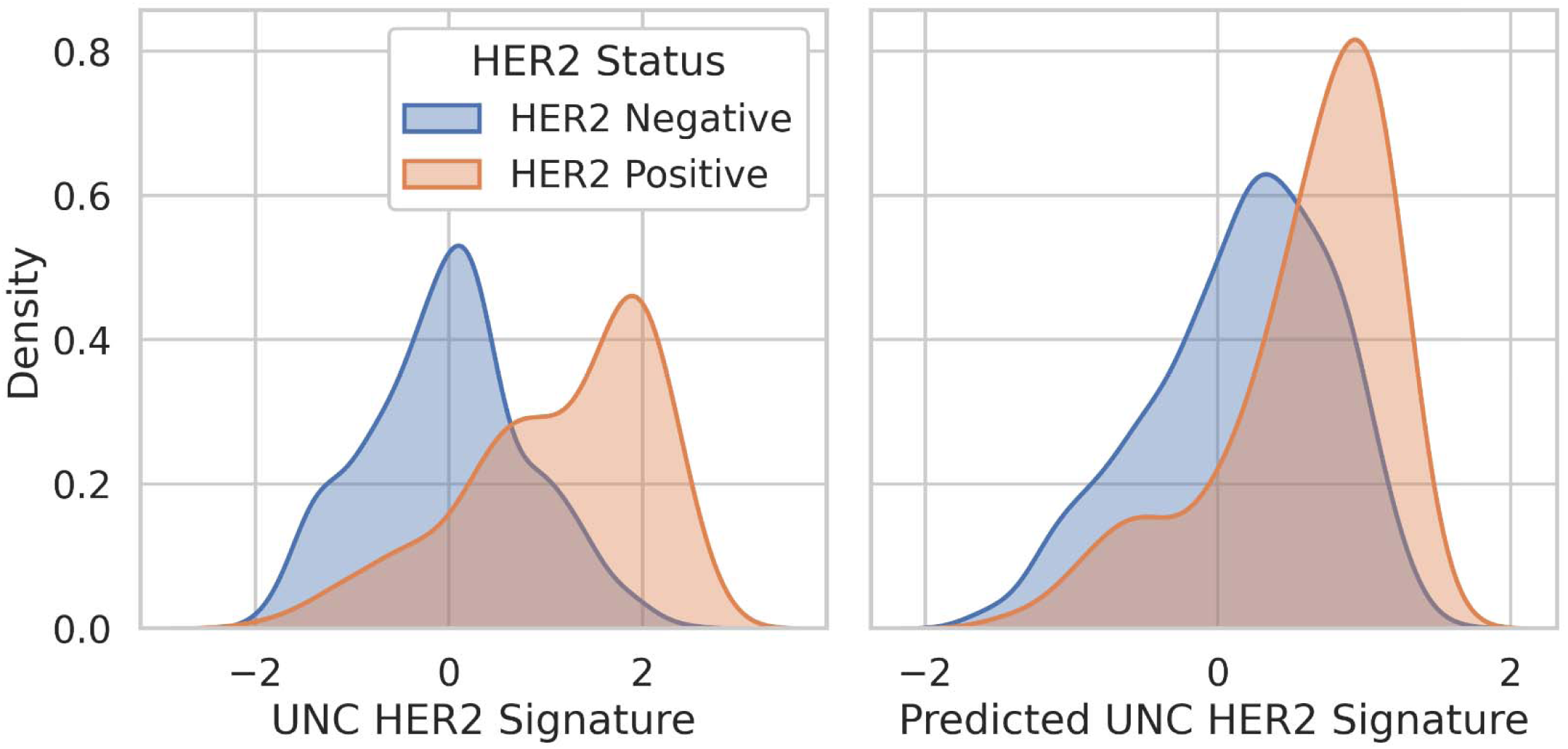
Distribution of Gene Signature Valus and Histology Signatures Predictions for a HER2 signature in The Cancer Genome Atlas. Shown are kernel density estimates of the distribution of both the true UNC HER2 signature (PMID 19204204) on the left, as well as the predicted values for this signature on the right, calculated on n = 783 HER2-negative and n = 154 HER2-positive cases in The Cancer Genome Atlas. The histology predictions are taken in aggregate from the held out test sets over five fold cross validation in this cohort. The ground truth gene signature more accurately separates HER2-positive and HER2-negative cases than the histology derived signature. **HER2**, human epidermal growth factor receptor 2; **UNC**, University of North Carolina.

**Extended Data Figure 9.**
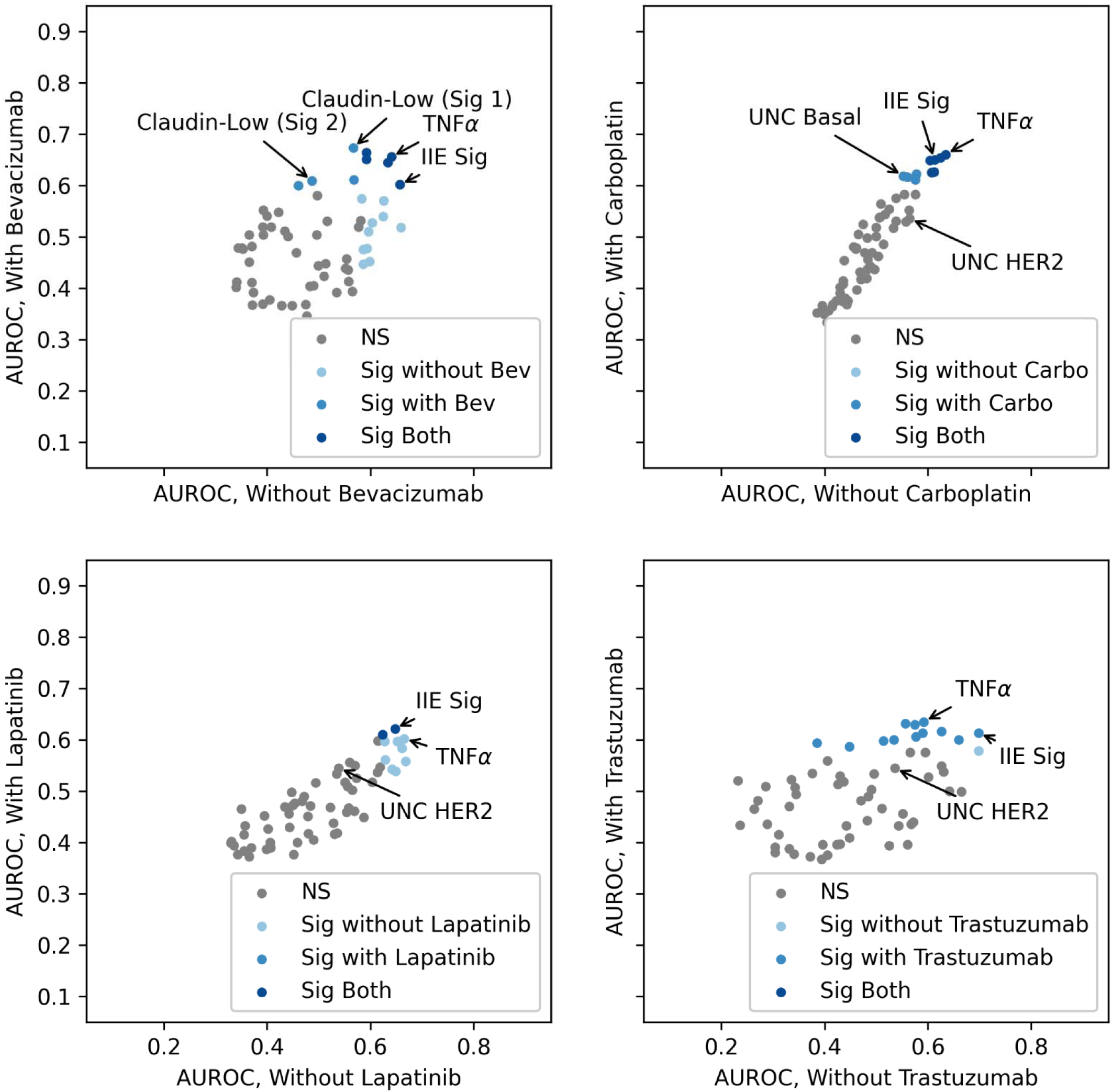
Histology-Derived Gene Expression Signatures can Predict Response **to Specific Treatments.** Gene expression signatures inferred from histology were evaluated for treatment-specific predictive power in the randomized CALGB 40603 and CALGB 40601 trials. Each plot compares AUROCs for predicting pathologic complete response (pCR) in patients receiving or not receiving a specific treatment within the trial. In CALGB 40603 (top row), two claudin-related signatures (from Herschkowitz et al, Genome Biology 2007) predicted response in the bevacizumab arm, but not with chemotherapy alone. Similarly, a basal-like signature was predictive of response to carboplatin, indicating treatment-specific sensitivity captured by histology-inferred gene signatures – although the generally linear association between response to treatment with or without carboplatin may indicate the lack of histology specific features for carboplatin response. In contrast, in CALGB 40601 (bottom row), no gene expression signatures showed strong differential performance across treatment arms including trastuzumab or lapatinib. As these assessments are considered exploratory, false discovery correction was not performed. **AUROC**, area under the receiver operating characteristic curve; **Bev**, bevacizumab; **CALGB**, Cancer and Leukemia Group B; **Carbo**, carboplatin; **HER2**, human epidermal growth factor receptor 2; **GES**, gene expression signature; **AUROC**, area under the receiver operating characteristic curve; **pCR**, pathological complete response; **UNC**, University of North Carolina. **TNF**α, tumor necrosis factor alpha.

**Extended Data Figure 10.**
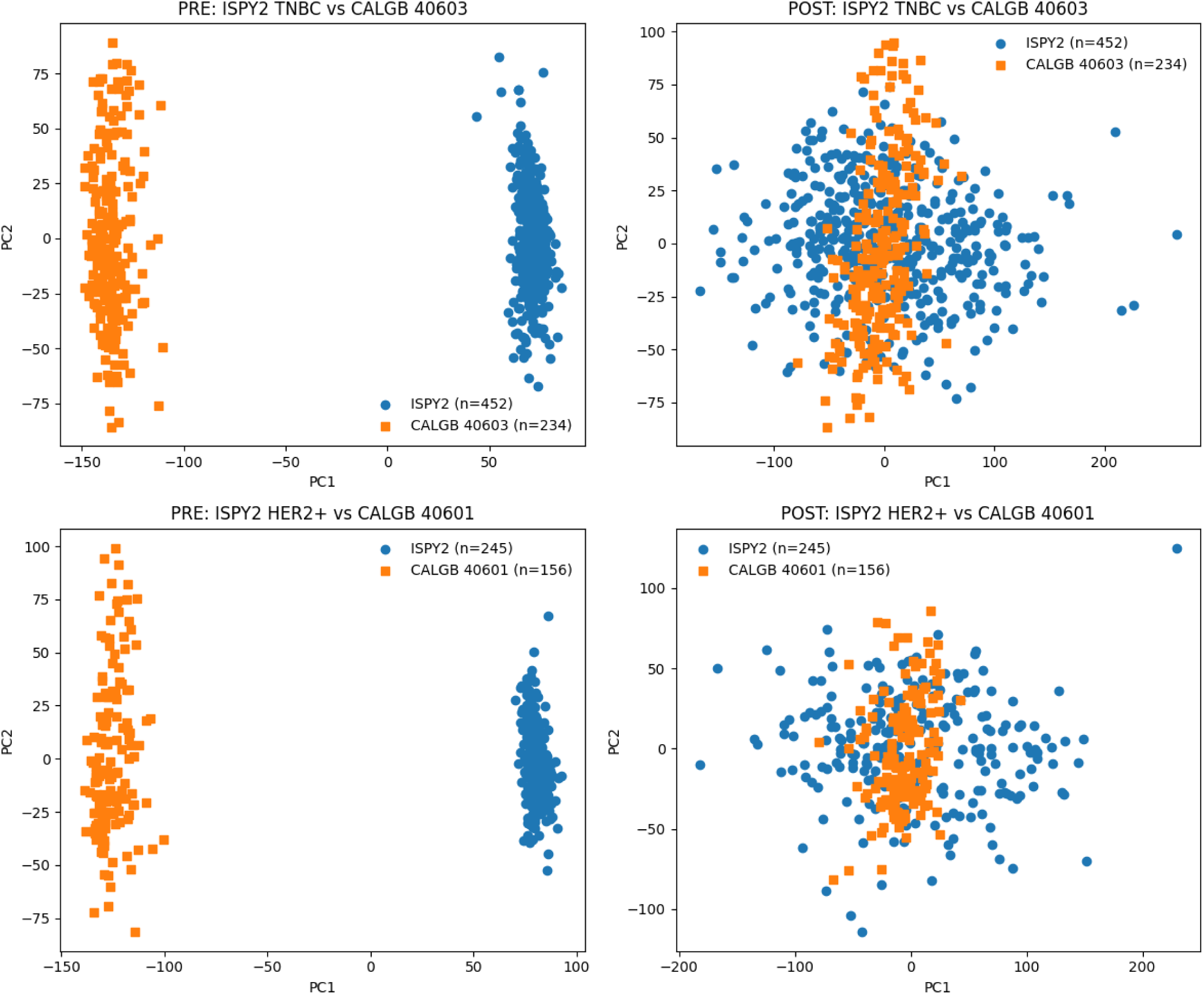
Batch Correction of Gene Expression Profiles Across Training and **Validation Cohorts.** Shown are scatter plots of the first two components on principle component analysis, demonstrating the clear batch related effect before and after batch correction. The top figures illustrate the TNBC cohort of I-SPY2, and the CALGB 40603 cohort (all TNBC), which was used to fit the batch correction model. The bottom figures demonstrate that this approach also harmonized the HER2+ cohort of I-SPY2 with the HER2+ CALGB 40601 without additional correction.

**Supplemental Table 1.** Correlation between Predicted and True Gene Expression Signatures Across Foundational Model / Feature Aggregation Methods. Models for 775 gene expression signatures were trained and evaluated with five-fold cross validation in TCGA-BRCA (n = 1052). Median and intraquartile range of Pearson correlation coefficient for true signature derived from RNAseq versus predicted signature value is reported across combinations of foundational model and feature aggregation architecture. The best performance was seen with the UNI foundational model and a variant of the Bistro Transformer architecture, which is subsequently used for all bulk gene expression and gene expression signature prediction tasks. Abbreviations: BRCA, breast invasive carcinoma; MIL, multiple-instance learning; RNAseq, RNA sequencing; TCGA, The Cancer Genome Atlas; IQR, interquartile range.

| Foundational Model | Feature Aggregation | Pearson r, median (IQR) |
| --- | --- | --- |
| CTransPath | Attention MIL | 0.505 (0.385 - 0.591) |
| CTransPath | Bistro Transformer | 0.523 (0.398 - 0.609) |
| CTransPath | TransMIL | 0.512 (0.390 - 0.597) |
| HistoSSL | Attention MIL | 0.501 (0.391 - 0.575) |
| HistoSSL | Bistro Transformer | 0.521 (0.424 - 0.594) |
| HistoSSL | TransMIL | 0.511 (0.413 - 0.587) |
| UNI | Attention MIL | 0.537 (0.417 - 0.613) |
| UNI | Bistro Transformer | 0.545 (0.428 - 0.620) |
| UNI | TransMIL | 0.538 (0.422 - 0.620) |

**Supplemental Table 2.** Comparative Accuracy of Optimized Gene Expression Prediction with TIGER versus Previous Methods. Gene expression models were trained and tested with internal five-fold cross validation in TCGA for six cancer subtypes, and externally tested in CPTAC, as well as the I-SPY2, CALGB 40601, and CALGB 40603 trials for the breast cancer cohort. Our optimized approach to gene expression prediction, TIGER, is compared to other previous models (SEQUOIA and DeepPT). Performance is measured as the average correlation coefficient for the top 1,000 predicted genes, with the top 1,000 genes identified separately for each model / cohort. Abbreviations: BRCA, breast invasive carcinoma; CALGB, Cancer and Leukemia Group B; COAD, colon adenocarcinoma; CPTAC, Clinical Proteomic Tumor Analysis Consortium; DeepPT, Deep learning-based Prediction of Transcriptomics; GBM, glioblastoma multiforme; HNSC, head and neck squamous cell carcinoma; I-SPY2, Investigation of Serial Studies to Predict Your Therapeutic Response with Imaging and Molecular Analysis 2; LUAD, lung adenocarcinoma; LUSC, lung squamous cell carcinoma; SEQUOIA, Spatially Enabled Quantification Of Individualized Alterations; TCGA, The Cancer Genome Atlas; TIGER, hIstology-driven Gene Expression Regressor.

| Subtype | Cohort | N | Comparator | Mean, Top 1000 | Mean, Top | T stat | P value |
| --- | --- | --- | --- | --- | --- | --- | --- |

**Supplemental Table 2. Comparative Accuracy of Optimized Gene Expression Prediction with TIGER versus Previous Methods.**
|  |  |  |  | Gene<br>Correlation,<br>TIGER | 1000 Gene<br>Correlation,<br>Comparator |  |  |
| --- | --- | --- | --- | --- | --- | --- | --- |
| BRCA | TCGA | 913 | SEQUOIA | 0.605 | 0.591 | 7.546953 | 6.73E-14 |
| BRCA | TCGA | 913 | DeepPT | 0.605 | 0.581 | 12.76048 | 6.62E-36 |
| BRCA | CPTAC | 88 | SEQUOIA | 0.647 | 0.637 | 5.305319 | 1.25E-07 |
| BRCA | CPTAC | 88 | DeepPT | 0.647 | 0.622 | 11.42955 | 2.73E-29 |
| BRCA | I-SPY | 230 | SEQUOIA | 0.509 | 0.470 | 15.22143 | 1.69E-49 |
| BRCA | I-SPY | 230 | DeepPT | 0.509 | 0.430 | 30.56047 | 9.20E-168 |
| BRCA | CALGB<br>40601 | 47 | SEQUOIA | 0.607 | 0.583 | 11.69545 | 1.32E-30 |
| BRCA | CALGB<br>40601 | 47 | DeepPT | 0.607 | 0.579 | 12.57147 | 6.88E-35 |
| BRCA | CALGB<br>40603 | 155 | SEQUOIA | 0.427 | 0.423 | 1.883706 | 0.059753 |
| BRCA | CALGB<br>40603 | 155 | DeepPT | 0.427 | 0.419 | 3.755977 | 0.000178 |
| COAD | TCGA | 421 | SEQUOIA | 0.552 | 0.548 | 1.419184 | 0.156002 |
| COAD | TCGA | 421 | DeepPT | 0.552 | 0.543 | 3.224002 | 0.001286 |
| COAD | CPTAC | 95 | SEQUOIA | 0.490 | 0.469 | 11.95477 | 7.39E-32 |
| COAD | CPTAC | 95 | DeepPT | 0.490 | 0.466 | 12.62104 | 3.82E-35 |
| GBM | TCGA | 94 | SEQUOIA | 0.106 | -0.010 | 6.28603 | 3.99E-10 |
| GBM | TCGA | 94 | DeepPT | 0.106 | 0.039 | 3.684352 | 0.000235 |
| GBM | CPTAC | 121 | SEQUOIA | 0.099 | 0.031 | 6.30979 | 3.61E-10 |
| GBM | CPTAC | 121 | DeepPT | 0.099 | 0.069 | 2.823581 | 0.004807 |
| HNSC | TCGA | 322 | SEQUOIA | 0.515 | 0.512 | 1.174023 | 0.240527 |
| HNSC | TCGA | 322 | DeepPT | 0.515 | 0.471 | 19.04837 | 7.15E-74 |
| HNSC | CPTAC | 81 | SEQUOIA | 0.436 | 0.456 | -9.15778 | 1.28E-19 |
| HNSC | CPTAC | 81 | DeepPT | 0.436 | 0.446 | -4.42691 | 1.01E-05 |
| LUSC | TCGA | 467 | SEQUOIA | 0.483 | 0.477 | 4.408265 | 1.10E-05 |
| LUSC | TCGA | 467 | DeepPT | 0.483 | 0.455 | 17.95021 | 6.81E-67 |
| LUSC | CPTAC | 89 | SEQUOIA | 0.565 | 0.541 | 12.54028 | 9.40E-35 |
| LUSC | CPTAC | 89 | DeepPT | 0.565 | 0.531 | 17.14773 | 1.87E-61 |
| LUAD | TCGA | 228 | SEQUOIA | 0.478 | 0.449 | 5.867627 | 5.20E-09 |
| LUAD | TCGA | 228 | DeepPT | 0.478 | 0.411 | 10.92905 | 5.18E-27 |
| LUAD | CPTAC | 174 | SEQUOIA | 0.517 | 0.453 | 16.93112 | 3.74E-60 |
| LUAD | CPTAC | 174 | DeepPT | 0.517 | 0.433 | 21.48927 | 4.19E-92 |

**Supplemental Table 3:** Description of Baseline Demographics of Cohorts Included in Analysis for Response Prediction. Abbreviations: CALGB, Cancer and Leukemia Group B; I-SPY, Investigation of Serial Studies to Predict Your Therapeutic Response with Imaging and Molecular Analysis; SD, standard deviation; T, tumor size staging category; N, nodal involvement staging category; M, presence of distant metastasis; HER2, human epidermal growth factor receptor 2; HR, hormone receptor; pCR, pathological complete response.

|  |  | **Overall** | **CALGB** | **I-SPY** | **UChicago** | **Yale** |
| --- | --- | --- | --- | --- | --- | --- |
| **n** |  | 1940 | 589 | 1115 | 195 | 41 |
| **Age, mean (SD)** |  | 49.0 (11.6) | 49.1 (10.5) | 48.3 (11.8) | 52.5 (13.5) | 50.5 (10.7) |
| **T, n (%)** | **T0** | 5 (0.3) | 4 (0.7) | 1 (0.1) |  |  |
|  | **T1** | 113 (5.8) | 33 (5.6) | 32 (2.9) | 36 (18.5) | 12 (29.3) |
|  | **T2** | 1199 (61.8) | 345 (58.6) | 720 (64.6) | 111 (56.9) | 23 (56.1) |
|  | **T3** | 497 (25.6) | 150 (25.5) | 302 (27.1) | 39 (20.0) | 6 (14.6) |
|  | **T4** | 63 (3.2) | 17 (2.9) | 37 (3.3) | 9 (4.6) |  |
|  | **Unknown** | 63 (3.2) | 40 (6.8) | 23 (2.1) |  |  |
| **N, n (%)** | **N0** | 826 (42.6) | 255 (43.3) | 471 (42.2) | 80 (41.0) | 20 (48.8) |
|  | **N1** | 835 (43.0) | 226 (38.4) | 502 (45.0) | 89 (45.6) | 18 (43.9) |
|  | **N2** | 116 (6.0) | 45 (7.6) | 52 (4.7) | 19 (9.7) |  |
|  | **N3** | 76 (3.9) | 9 (1.5) | 58 (5.2) | 6 (3.1) | 3 (7.3) |
|  | **Unknown** | 87 (4.5) | 54 (9.2) | 32 (2.9) | 1 (0.5) |  |
| **M, n (%)** | **M0** | 1791 (92.3) | 589 (100.0) | 969 (86.9) | 192 (98.5) | 41 (100.0) |
|  | **M1** | 3 (0.2) |  | 1 (0.1) | 2 (1.0) |  |
|  | **Unknown** | 146 (7.5) |  | 145 (13.0) | 1 (0.5) |  |
| **HER2, n (%)** | **Negative** | 1422 (73.3) | 359 (61.0) | 886 (79.5) | 136 (69.7) | 41 (100.0) |
|  | **Positive** | 517 (26.6) | 230 (39.0) | 228 (20.4) | 59 (30.3) |  |
|  | **Unknown** | 1 (0.1) |  | 1 (0.1) |  |  |
| **HR, n (%)** | **Negative** | 1031 (53.1) | 453 (76.9) | 443 (39.7) | 94 (48.2) | 41 (100.0) |
|  | **Positive** | 907 (46.8) | 135 (22.9) | 671 (60.2) | 101 (51.8) |  |
|  | **Unknown** | 2 (0.1) | 1 (0.2) | 1 (0.1) |  |  |
| **Grade, n (%)** | **1** | 34 (1.8) | 15 (2.5) | 16 (1.4) | 3 (1.5) |  |
|  | **2** | 377 (19.4) | 100 (17.0) | 243 (21.8) | 34 (17.4) |  |
|  | **3** | 1152 (59.4) | 429 (72.8) | 565 (50.7) | 158 (81.0) |  |
|  | **Unknown** | 377 (19.4) | 45 (7.6) | 291 (26.1) |  | 41 (100.0) |
| **pCR, n (%)** | **No** | 1211 (62.4) | 322 (54.7) | 742 (66.5) | 124 (63.6) | 23 (56.1) |
|  | **Yes** | 729 (37.6) | 267 (45.3) | 373 (33.5) | 71 (36.4) | 18 (43.9) |

**Supplemental Table 4:** Treatment Regimens Received in Patients Included in the Analysis. Abbreviations: AC, doxorubicin and cyclophosphamide; CALGB, Cancer and Leukemia Group B; I-SPY, Investigation of Serial Studies to Predict Your Therapeutic Response with Imaging and Molecular Analysis.

|  | **CALGB** | **I-SPY** | **UChicago** | **Yale** |
| --- | --- | --- | --- | --- |
|  | 589 | 1115 | 195 | 41 |
| **Paclitaxel + Lapatinib** | 49 (8.3) |  |  |  |
| **Paclitaxel + Lapatinib + Trastuzumab** | 95 (16.1) |  |  |  |
| **Paclitaxel + Bevacizumab → AC** | 84 (14.3) |  |  |  |
| **Paclitaxel + Bevacizumab + Carboplatin → AC** | 83 (14.1) |  |  |  |
| **Paclitaxel + Carboplatin → AC** | 95 (16.1) |  |  |  |
| **Paclitaxel + Trastuzumab** | 86 (14.6) |  |  |  |
| **Paclitaxel → AC** | 97 (16.5) | 343 (30.8) | 103 (52.8) |  |
| **Oral Paclitaxel + Encequidar + Dostarlimab + Carboplatin + Trastuzumab → AC** |  | 5 (0.4) |  |  |
| **Oral Paclitaxel + Encequidar + Dostarlimab + Carboplatin → AC** |  | 96 (8.6) |  |  |
| **Oral Paclitaxel + Encequidar + Dostarlimab + Trastuzumab → AC** |  | 18 (1.6) |  |  |
| **Oral Paclitaxel + Encequidar + Dostarlimab → AC** |  | 95 (8.5) |  |  |
| **Paclitaxel + Cemiplimab + REGN3767 → AC** |  | 71 (6.4) |  |  |
| **Paclitaxel + Cemiplimab → AC** |  | 50 (4.5) |  |  |
| **Paclitaxel + Pembrolizumab 8-Cycle + AC** |  | 90 (8.1) |  |  |
| **Paclitaxel + Pembrolizumab → AC** |  | 67 (6.0) |  |  |
| **SD-101 + Pembrolizumab → AC** |  | 74 (6.6) |  |  |
| **Paclitaxel + Trastuzumab → AC** |  | 27 (2.4) | 5 (2.6) |  |
| **Paclitaxel + Trastuzumab + Pertuzumab → AC** |  | 179 (16.1) | 17 (8.7) |  |
| **AC** |  |  | 2 (1.0) |  |
| **Other** |  |  | 19 (9.7) |  |
| **Paclitaxel + Carboplatin** |  |  | 6 (3.1) |  |
| **Paclitaxel + Carboplatin + Trastuzumab** |  |  | 12 (6.2) |  |
| **Paclitaxel + Carboplatin + Trastuzumab → AC** |  |  | 3 (1.5) |  |
| **Paclitaxel + Carboplatin + Trastuzumab + Pertuzumab** |  |  | 18 (9.2) |  |
| **Paclitaxel + Trastuzumab + Pertuzumab** |  |  | 3 (1.5) |  |
| **Durvalumab + nab-Paclitaxel → AC** |  |  |  | 41 (100.0) |

**Supplemental Table 5:** Set of 61 Signatures Included for Statistical Testing of Response Association. Short names of specific signatures as referenced throughout the manuscript are provided.

| Signature Reference | Signature Short Name |
| --- | --- |
| Pcorr_NKI70_Good_Correlation_Nature.2002_PMID.11823860 |  |
| Wound.Healing_Chang_PlosBiol.2004_PMID.14737219 |  |
| GHI_RS_Model_NJEM.2004_PMID.15591335 |  |
| MCF7.E2.repressed.genes_JCO.2006_PMID.16505416 |  |
| Scorr_IIE_Correlation_JCO.2006_PMID.16505416 | IIE |
| Scorr_IE_Correlation_JCO.2006_PMID.16505416 |  |
| MCF7.E2.induced.genes_JCO.2006_PMID.16505416 |  |
| Pcorr_IGS_Correlation_NJEM.2007_PMID.17229949 |  |
| Pcorr_Dasatinib_R_Correlation_Cancer.Res.2007_PMID.17332353 |  |
| Pcorr_Dasatinib_L_Correlation_Cancer.Res.2007_PMID.17332353 |  |
| Shipitsin_CD44_A_Cancer.Cell.2007_PMID.17349583 |  |
| Claudin_Low_Genome.Biol.2007_PMID.17493263 |  |
| MM_DMBAwnt_1pFDR_UP_Genome.Biology.2007_PMID.17493263 |  |
| Claudin_High_Genome.Biol.2007_PMID.17493263 | Claudin-Low (Sig 1) |
| MM_Normal_1pFDR_UP_Genome.Biology.2007_PMID.17493263 |  |
| GSEA_GP5_MYC_targets_TERT.r.0.922_PerouLab_MM_Myc_1pFDR_UP_Genome_Biology_2007_PMID.17493263 |  |
| MM_Potluck_1pFDR_UP_Genome.Biology.2007_PMID.17493263 | Claudin-Low (Sig 2) |
| Fibromatosis_Lab.Invest.2008_PMID.18414401 |  |
| UNC_Scorr_LumB_Correlation_JCO.2009_PMID.19204204 |  |
| UNC_Scorr_Her2_Correlation_JCO.2009_PMID.19204204 | UNC HER2 |
| UNC_Scorr_Norm_Correlation_JCO.2009_PMID.19204204 |  |
| UNC_Scorr_Basal_Correlation_JCO.2009_PMID.19204204 | UNC Basal |
| VEGF_13genes_BMC.Med.2009_PMID.19291283 |  |
| Glycolysis_Signature_BMC.Med.2009_PMID.19291283 |  |
| Luminal_Progenitor_Down_Nat.Med.2009_PMID.19648928 |  |
| Mature_LuminaUp_Nat.Med.2009_PMID.19648928 |  |
| Duke_Module22_tnfa_Mike_PMID.20335537 | TNFα Signature |
| Duke_Module07_glucosedepletion_Mike_PMID.20335537 |  |
| HS_Green14_BMC.Med.Genomics.2011_PMID.21214954 |  |
| MM_Red23_BMC.Med.Genomics.2011_PMID.21214954 |  |
| HS_Green13_BMC.Med.Genomics.2011_PMID.21214954 |  |
| MUnknown_31_BMC.Med.Genomics.2011_PMID.21214954 |  |
| MM_Green4_BMC.Med.Genomics.2011_PMID.21214954 |  |
| HS_Red22_BMC.Med.Genomics.2011_PMID.21214954 |  |
| HS_Green20_BMC.Med.Genomics.2011_PMID.21214954 |  |
| MUnknown_24_BMC.Med.Genomics.2011_PMID.21214954 |  |
| MUnknown_18_BMC.Med.Genomics.2011_PMID.21214954 |  |
| HS_Red10_BMC.Med.Genomics.2011_PMID.21214954 |  |
| HS_Green12_BMC.Med.Genomics.2011_PMID.21214954 |  |
| MUnknown_15_BMC.Med.Genomics.2011_PMID.21214954 |  |
| MM_Green5_BMC.Med.Genomics.2011_PMID.21214954 |  |
| Unknown_9_BMC.Med.Genomics.2011_PMID.21214954 |  |
| HS_Green21_BMC.Med.Genomics.2011_PMID.21214954 |  |
| S100A9_A8_BMC.Med.Genomics.2011_PMID.21214954 |  |
| HS_Red11_BMC.Med.Genomics.2011_PMID.21214954 |  |
| TNBC_Clinically_Relevant_Good.230_BCR.2011_PMID.21978456 |  |
| TNBC_Clinically_Relevant_Poor.26_BCR.2011_PMID.21978456 |  |
| Wahl_fMasC_Signature_Cell.Stem.Cell.2012_PMID.22305568 |  |
| Pcorr_bronchioid_PLOS.2012_PMID.22590557 |  |
| Pcorr_squamoid_PLOS.2012_PMID.22590557 |  |
| MM_Neu.2012_Genome.Biol.2013_PMID.24220145 |  |
| Lim2009_MaSC_Adam_PMID.25575446 |  |
| aMaSC_Lim09_BCR.2015_PMID.25575446 |  |
| Melanoma_Scorr_MITF.low_Cell.2015_PMID.26091043 |  |
| Melanoma_Scorr_Keratin_Cell.2015_PMID.26091043 |  |
| Epithelial_Tubule_Formation_J.Pathol.2017_PMID.27861902 |  |
| Apocrine_Features_J.Pathol.2017_PMID.27861902 |  |
| PR_Isoform_Ratio_Up_in_PRA_H_JNCI.2017_PMID.28376177 |  |
| PARP_sensitivity_MDACC_NPJ.Syst.Biol.Appl._2017_PMID.28649435 |  |
| Scorr_EMAT2_Correlation_BCR.2020_PMID.32641077 |  |
| Scorr_EMAT3_Correlation_BCR.2020_PMID.32641077 |  |

**Supplemental Table 6:** Comparison of TILs and IIE Signature Prediction for Assessment of Response to Therapy. Abbreviations: AUROC, area under the receiver operating characteristic curve; CALGB, Cancer and Leukemia Group B; CI, confidence interval; TIL, Tumor infiltrating lymphocyte

| **Cohort** | **N Patients** | **AUROC TILs (95% CI)** | **AUROC IIE Signature (95% CI)** | **p-value** |
| --- | --- | --- | --- | --- |
| Overall | 516 | 0.62 (0.57-0.67) | 0.68 (0.63-0.72) | 0.005 |
| CALGB 40603 | 156 | 0.64 (0.55-0.73) | 0.65 (0.56-0.73) | 0.79 |
| CALGB 40601 | 129 | 0.57 (0.46-0.67) | 0.60 (0.50-0.70) | 0.448 |
| UChicago | 190 | 0.60 (0.52-0.69) | 0.72 (0.65-0.80) | 0.003 |
| Yale | 41 | 0.66 (0.49-0.83) | 0.74 (0.58-0.89) | 0.306 |

**Supplemental Table 7:**
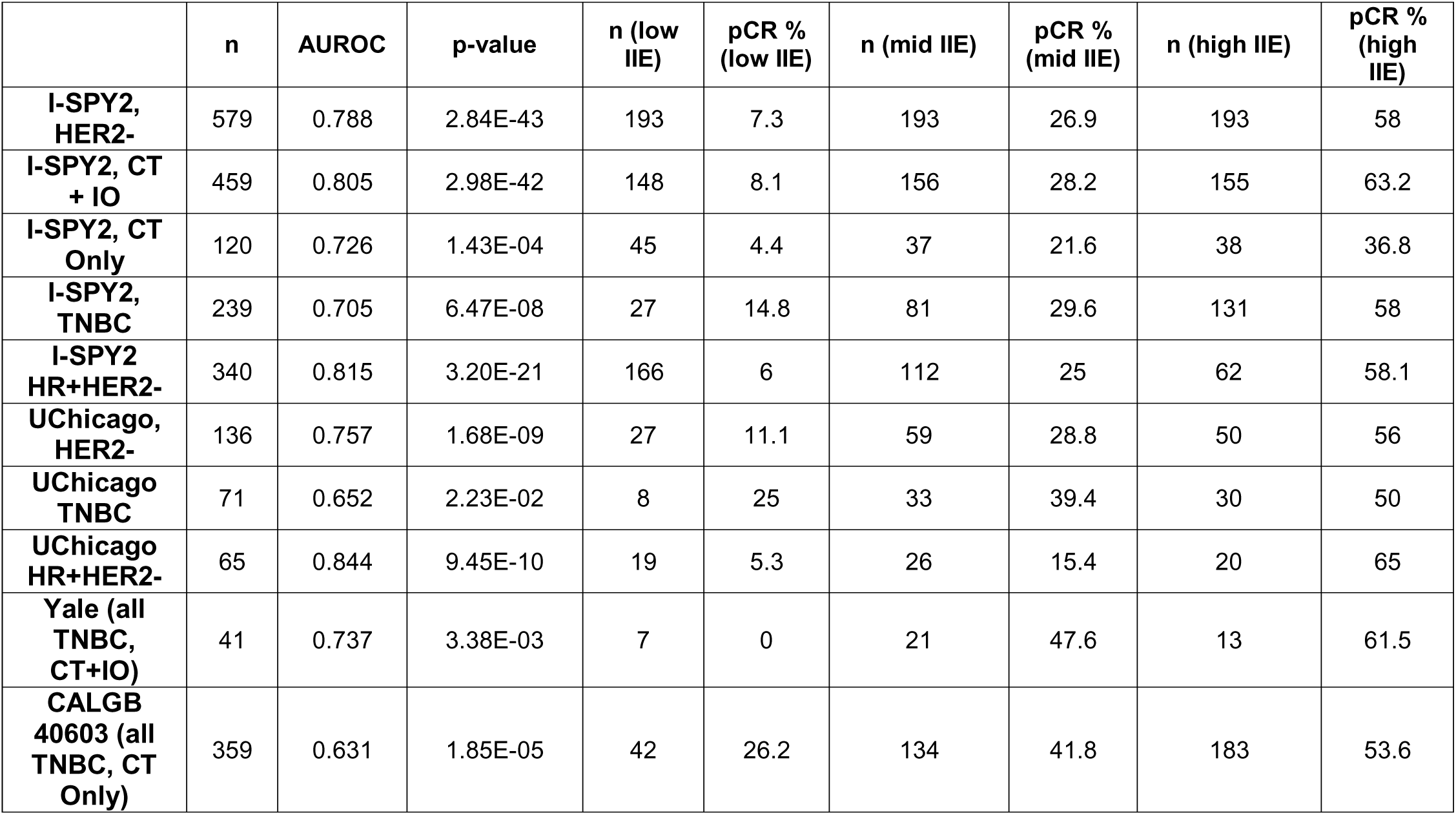
Accuracy for pCR Prediction with Predicted IIE Signature Across Subgroups of Patients with HER2- Breast Cancer. Abbreviations: AUROC, area under the receiver operating characteristic curve; CALGB, Cancer and Leukemia Group B; CT, chemotherapy; HER2, human epidermal growth factor receptor 2; HR, hormone receptor; IO, immunotherapy; I-SPY, Investigation of Serial Studies to Predict Your Therapeutic Response with Imaging and Molecular Analysis; pCR, pathological complete response; TNBC, triple-negative breast cancer.

**Supplemental Table 8:** Accuracy for RCB 0-1 (versus 2-3) Prediction with Predicted IIE Signature Across Subgroups of Patients with HER2- Breast Cancer. Abbreviations: AUROC, area under the receiver operating characteristic curve; CALGB, Cancer and Leukemia Group B; CT, chemotherapy; HER2, human epidermal growth factor receptor 2; HR, hormone receptor; IO, immunotherapy; I-SPY, Investigation of Serial Studies to Predict Your Therapeutic Response with Imaging and Molecular Analysis; RCB, residual cancer burden; TNBC, triple-negative breast cancer.

|  | n | AUROC | p-value | n (low IIE) | RCB 0-1 % (low IIE) | n (mid IIE) | RCB 0-1 % (mid IIE) | n (high IIE) | RCB 0-1 % (high IIE) |
| --- | --- | --- | --- | --- | --- | --- | --- | --- | --- |
| **I-SPY2, HER2-** | 684 | 0.76 | 4.02E-37 | 209 | 25.4 | 239 | 51.9 | 206 | 78.6 |
| **I-SPY2, CT + IO** | 459 | 0.795 | 7.80E-38 | 138 | 20.3 | 147 | 46.3 | 151 | 82.1 |
| **I-SPY2, CT Only** | 120 | 0.674 | 6.23E-04 | 44 | 31.8 | 34 | 38.2 | 36 | 63.9 |
| **I-SPY2, TNBC** | 239 | 0.683 | 8.46E-06 | 26 | 42.3 | 75 | 50.7 | 125 | 79.2 |
| **I-SPY2 HR+HER2-** | 340 | 0.771 | 3.00E-23 | 156 | 19.9 | 106 | 40.6 | 62 | 77.4 |
| **Yale (all TNBC, CT+IO)** | 41 | 0.706 | 2.78E-02 | 7 | 14.3 | 20 | 65 | 13 | 69.2 |
| **CALGB 40603 (all TNBC, CT Only)** | 359 | 0.628 | 1.62E-05 | 42 | 45.2 | 134 | 50.7 | 183 | 63.9 |

**Supplemental Table 9:** Accuracy for pCR Prediction with Predicted HER2 Signature Across Subgroups of Patients with HER2- Breast Cancer Treated with Chemotherapy Alone versus Chemotherapy with Immunotherapy. Abbreviations: AUROC, area under the receiver operating characteristic curve; CALGB, Cancer and Leukemia Group B; CT, chemotherapy; HER2, human epidermal growth factor receptor 2; IO, immunotherapy; I-SPY, Investigation of Serial Studies to Predict Your Therapeutic Response with Imaging and Molecular Analysis; pCR, pathologic complete response.

|  | n | AUROC | p-value | n (low<br>HER2) | pCR %<br>(low<br>HER2) | n (mid<br>HER2) | pCR %<br>(mid<br>HER2) | n (high<br>HER2) | pCR %<br>(high<br>HER2) |
| --- | --- | --- | --- | --- | --- | --- | --- | --- | --- |
| I-SPY2, CT<br>+ IO | 459 | 0.661 | 1.59E-08 | 165 | 13.9 | 150 | 44.7 | 144 | 44.4 |
| Yale (CT +<br>IO) | 41 | 0.681 | 4.16E-02 | 5 | 0 | 12 | 58.3 | 24 | 45.8 |
| I-SPY2, CT<br>Only | 120 | 0.547 | 7.63E-01 | 28 | 14.3 | 43 | 23.3 | 49 | 20.4 |
| UChicago,<br>HER2- (CT<br>only) | 136 | 0.526 | 6.14E-01 | 40 | 35 | 46 | 30.4 | 50 | 40 |
| CALGB<br>40603 (CT<br>Only) | 359 | 0.55 | 1.09E-01 | 70 | 37.1 | 96 | 42.7 | 193 | 50.8 |

**Supplemental Table 10:**
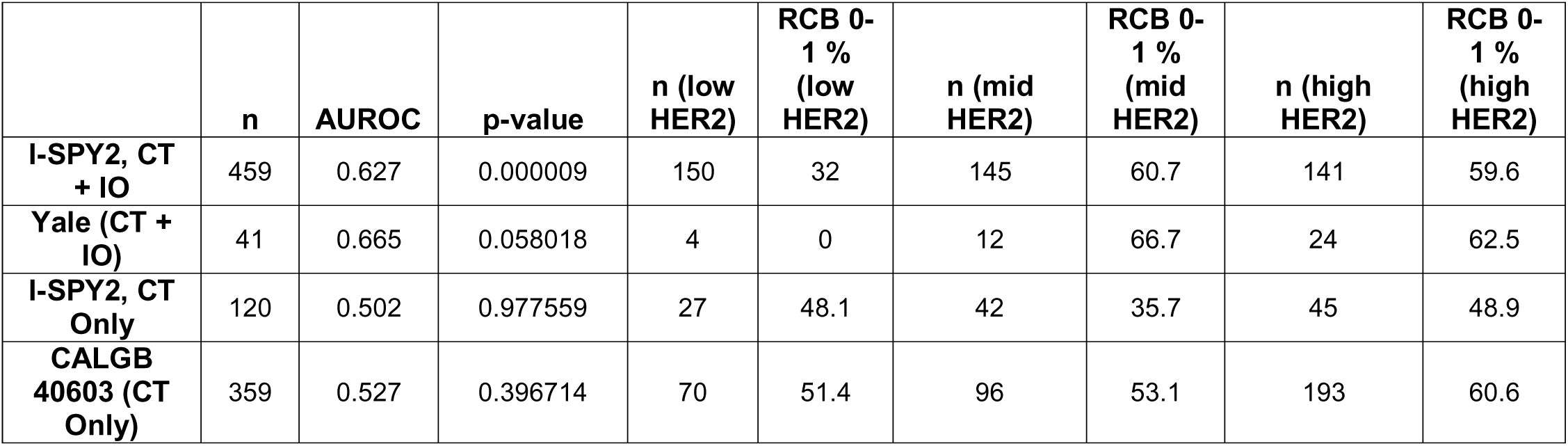
Accuracy for RCB 0-1 (versus 2-3) with Predicted HER2 Signature Across Subgroups of Patients with HER2- Breast Cancer Treated with Chemotherapy Alone versus Chemotherapy with Immunotherapy. Abbreviations: AUROC, area under the receiver operating characteristic curve; CALGB, Cancer and Leukemia Group B; CT, chemotherapy; HER2, human epidermal growth factor receptor 2; IO, immunotherapy; I-SPY, Investigation of Serial Studies to Predict Your Therapeutic Response with Imaging and Molecular Analysis; RCB, residual cancer burden.

**Supplemental Table 11:** Accuracy for pCR Prediction with Predicted TNFα Signature Across Subgroups of Patients with HER2+ Breast Cancer. Abbreviations: AUROC, area under the receiver operating characteristic curve; CALGB, Cancer and Leukemia Group B; HER2, human epidermal growth factor receptor 2; HR, hormone receptor; I-SPY, Investigation of Serial Studies to Predict Your Therapeutic Response with Imaging and Molecular Analysis; pCR, pathologic complete response; TNFα, tumor necrosis factor alpha.

|  | n | AUROC | p-value | n (low TNF) | pCR % (low TNF) | n (mid TNF) | pCR % (mid TNF) | n (high TNF) | pCR % (high TNF) |
| --- | --- | --- | --- | --- | --- | --- | --- | --- | --- |
| **I-SPY2, HER2+** | 105 | 0.668 | 0.0498 | 33 | 39.4 | 43 | 55.8 | 29 | 69 |
| **I-SPY2, HR-HER2+** | 31 | 0.546 | 0.9289 | 6 | 83.3 | 15 | 86.7 | 10 | 80 |
| **I-SPY2, HR+HER2+** | 74 | 0.665 | 0.0870 | 27 | 29.6 | 28 | 39.3 | 19 | 63.2 |
| **UChicago HER2+** | 59 | 0.748 | 0.0002 | 28 | 21.4 | 22 | 50 | 9 | 66.7 |
| **UChicago HR-HER2+** | 23 | 0.629 | 0.2935 | 8 | 37.5 | 11 | 54.5 | 4 | 50 |
| **UChicago HR+HER2+** | 36 | 0.767 | 0.0073 | 20 | 15 | 11 | 45.5 | 5 | 80 |
| **CALGB 40601** | 230 | 0.626 | 0.003158 | 93 | 33.3 | 73 | 47.9 | 64 | 56.2 |
| **CALGB 40601 HR-** | 94 | 0.47 | 1.333656 | 24 | 58.3 | 30 | 50 | 40 | 55 |
| **CALGB 40601 HR+** | 135 | 0.686 | 0.000878 | 69 | 24.6 | 42 | 45.2 | 24 | 58.3 |

**Supplemental Table 12:**
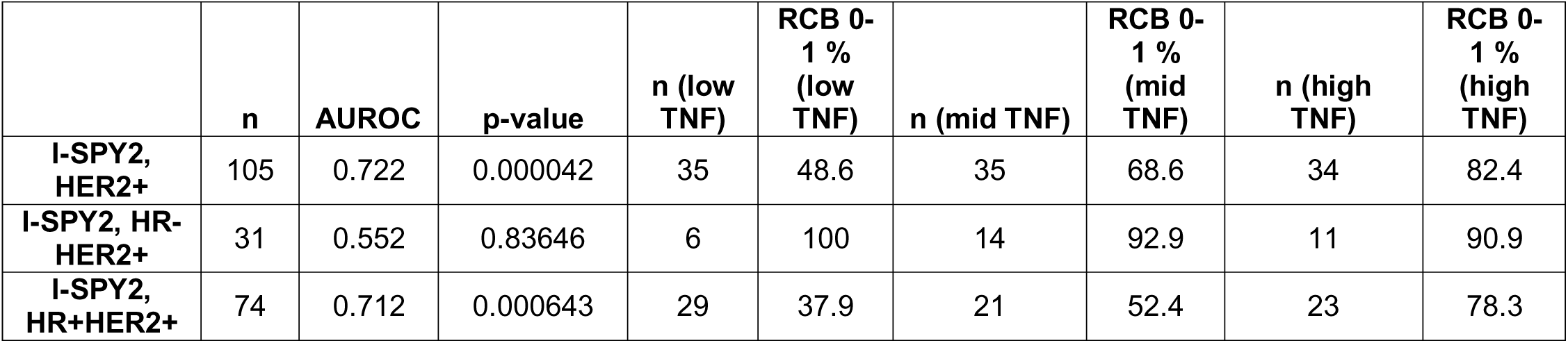
Accuracy for RCB 0-1 (versus 2-3) Prediction with Predicted TNFα Signature Across Subgroups of Patients with HER2+ Breast Cancer. Abbreviations: AUROC, area under the receiver operating characteristic curve; HER2, human epidermal growth factor receptor 2; HR, hormone receptor; I-SPY, Investigation of Serial Studies to Predict Your Therapeutic Response with Imaging and Molecular Analysis; RCB, residual cancer burden; TNFα, tumor necrosis factor alpha.

|  | n | AUROC | p-value | n (low TNF) | RCB 0-1 % (low TNF) | n (mid TNF) | RCB 0-1 % (mid TNF) | n (high TNF) | RCB 0-1 % (high TNF) |
| --- | --- | --- | --- | --- | --- | --- | --- | --- | --- |
| **I-SPY2, HER2+** | 105 | 0.722 | 0.000042 | 35 | 48.6 | 35 | 68.6 | 34 | 82.4 |
| **I-SPY2, HR-HER2+** | 31 | 0.552 | 0.83646 | 6 | 100 | 14 | 92.9 | 11 | 90.9 |
| **I-SPY2, HR+HER2+** | 74 | 0.712 | 0.000643 | 29 | 37.9 | 21 | 52.4 | 23 | 78.3 |

**Supplemental Table 13:** Prediction of Pathologic Complete Response from Pathology, RNA, or a Combination. Abbreviations: AUROC, area under the receiver operating characteristic curve; CI, confidence interval; HER2, human epidermal growth factor receptor 2; I-SPY, Investigation of Serial Studies to Predict Your Therapeutic Response with Imaging and Molecular Analysis; CALGB, Cancer and Leukemia Group B.

| Cohort | Modality | n | AUROC (95% CI) | p-value (vs RNA model) | p-value (vs combined model) |
| --- | --- | --- | --- | --- | --- |
| I-SPY2, HER2- | Path (IIE sig) | 238 | 0.701 (0.627–0.774) | 0.536 | 0.0393 |
|  | RNA (full) | 742 | 0.739 (0.701–0.778) | -- | -- |
|  | RNA (limited) | 238 | 0.726 (0.651–0.800) | -- | 0.221 |
|  | RNA + Path | 238 | **0.752 (0.684–0.820)** | 0.221 | -- |
| CALGB 40603 | Path (IIE sig) | 195 | 0.640 (0.562–0.717) | 0.199 | 0.0133 |
|  | RNA (full) | 234 | 0.663 (0.593–0.733) | -- | -- |
|  | RNA (limited) | 195 | 0.705 (0.632–0.778) | -- | 0.318 |
|  | RNA + Path | 195 | **0.730 (0.659–0.800)** | 0.318 | -- |
| I-SPY2 HER2+ | Path (IIE sig) | 70 | 0.756 (0.643–0.869) | 0.644 | 0.258 |
|  | RNA (full) | 245 | 0.743 (0.682–0.805) | -- | -- |
|  | RNA (limited) | 70 | 0.776 (0.665–0.887) | -- | 0.727 |
|  | RNA + Path | 70 | **0.785 (0.675–0.895)** | 0.727 | -- |
| CALGB 40601 | Path (IIE sig) | 146 | 0.571 (0.475–0.668) | 0.00571 | 0.00756 |
|  | RNA (full) | 156 | 0.686 (0.599–0.773) | -- | -- |
|  | RNA (limited) | 146 | **0.689 (0.599–0.778)** | -- | 0.0639 |
|  | RNA + Path | 146 | 0.637 (0.544–0.729) | 0.0639 | -- |

